# Estimating and explaining the spread of COVID-19 at the county level in the USA

**DOI:** 10.1101/2020.06.18.20134700

**Authors:** Anthony R. Ives, Claudio Bozzuto

## Abstract

The basic reproduction number, R_0_, determines the rate of spread of a communicable disease and therefore gives fundamental information needed to plan public health interventions. Using mortality records, we estimated the rate of spread of COVID-19 among 160 counties and county-aggregates in the USA. Here, we show that most of the high among-county variance is explainable by four factors (R^2^ = 0.70): the timing of outbreak, population size, population density, and spatial location. For predictions of future spread, population density and spatial location are important, and for the latter we show that SARS-CoV-2 strains containing the G614 mutation to the spike gene are associated with higher rates of spread. Finally, the high predictability of R_0_ allows extending estimates to all 3109 counties in the conterminous 48 states. The high variation of R_0_ argues for public health policies enacted at the county level for controlling COVID-19.

## Introduction

The basic reproduction number, R_0_, is the number of secondary infections produced per primary infection of a disease in a susceptible population, and it is a fundamental metric in epidemiology that gauges, among other factors, the initial rate of disease spread during an epidemic^1^. While R_0_ depends in part on the biological properties of the pathogen, it also depends on properties of the host population such as the contact rate between individuals^1,2^. Estimates of R_0_ are required for designing public health interventions for infectious diseases such as COVID-19: for example, R_0_ determines in large part the proportion of a population that must be vaccinated to control a disease^3,4^. Because R_0_ at the start of an epidemic measures the spread rate under “normal” conditions without interventions, these initial R_0_ values can inform policies to allow life to get “back to normal.”

The estimates of R_0_ before intervention determine the intensity with which public health interventions must be applied, and the risk and magnitude of potential resurgent outbreaks. In these contexts, R_0_ is a reference against which the success or failure of public interventions can be assessed. Using R_0_ estimates to design public health policies is predicated on the assumption that the R_0_ values at the start of the epidemic reflect properties of the infective agent and population, and therefore predict the potential rate of spread of the disease. Estimates of R_0_, however, might not predict future risks if (i) they are measured after public and private actions have been taken to reduce spread^5,6^, (ii) they are driven by stochastic events, such as super-spreading^7,8^, or (iii) they are driven by social or environmental conditions that are likely to change between the time of initial epidemic and the future time for which public health interventions are designed^9,10^. To address these potential limitations for using R_0_ to design public health policies and future risks of spread, we investigated possible underlying causes for variation in estimates of R_0_ among counties: if the causes are unlikely to change in the future, then so too are values of R_0_ unlikely to change.

Policies to manage for COVID-19 in the USA are set by a mix of jurisdictions from state to local levels. We estimated R_0_ at the county level both to match policymaking and to account for possibly large variation in R_0_ among counties. To estimate R_0_, we performed the analyses on the number of daily COVID-19 deaths^11^. We used death count rather than infection case reports, because we suspected the proportion of reported deaths due to COVID-19 is less sensitive to variation in testing rates and methods. We recognize that some deaths due to COVID-19 will go unreported (e.g., the growing evidence from “excess deaths”^12^) and that different counties and states may use different criteria for determining the cause of death as COVID-19. Nonetheless, due to the mathematical structure of our estimation procedure, unreported deaths due to COVID-19 and differences among counties in reporting criteria will have little effect on our estimates of R_0_; specifically, the estimates of R_0_ for a given county will not change provided the proportion of unreported deaths in a county does not change through time. We analyzed data for counties that had at least 100 reported cumulative deaths (Methods), and for other counties we aggregated data within the same state including deaths whose county was unknown. This led to 160 final time series representing counties in 39 states and the District of Columbia, of which 36 were aggregated at the state level. Some states, even after aggregating data from all counties, did not reach the 100-threshold of cumulative deaths, and therefore the spread rate for these states was not estimated.

We found high variance in the spread rate of COVID-19 among counties, most of which is explained by four factors: the timing of the county-level outbreak, population size, population density, and spatial location. Population density is likely an indicator of the average contact rate among people, and its explanatory power in the statistical model makes it an important predictor of future spread. Spatial location is also important, and we show that some of the effect of spatial location could be caused by differences among strains of SARS-CoV-2 that dominated in different parts of the USA. Using the statistical model, we estimated R_0_ at the county level for the entire conterminous USA, giving information to design public health policies for controlling COVID-19.

## Results

### Estimates of the spread rate

Before estimating R_0_, we first estimated the rate of spread of the virus-caused COVID-19 as the rate of increase of the daily death counts, *r*_0_. Although this approach is not typically used in epidemiological studies, it has the advantage of being statistically robust even when the data (death counts) are low, and it makes the minimum number of assumptions that could affect the estimates in unexpected ways (Supplementary Methods: Overview of Statistical Methods). We applied a time-varying autoregressive state-space model to each time series of death counts^13^. In contrast to other models of COVID-19 epidemics^14,15^, we do not incorporate the transmission process and the daily time course of transmission, but instead we estimate the time-varying exponential change in the number of deaths per day, *r*(*t*). Detailed simulation analyses (Supplementary Methods: Simulation model) showed that estimates of *r*(*t*) generally lagged behind the true values. Therefore, we analyzed the time series in forward and reverse directions, and averaged to get the estimates of *r*_0_ at the start of the time series (Supplementary Fig. S1); this approach counterbalances the lag in the forward direction with the lag in the backwards direction, therefore reducing the lag effect. The model was fit accounting for greater uncertainty when mortality counts were low, and confidence intervals of the estimates were obtained from parametric bootstrapping, which is the most robust approach when counts are low. Thus, our strategy was to use a parsimonious model to give robust estimates of *r*_0_ even for counties that had experienced relatively few deaths, and then calculate R_0_ from *r*_0_ after the fitting process using well-established methods^16^.

Our *r*_0_ estimates ranged from close to zero for several counties to 0.33 for New York City (five boroughs); the latter implies that the number of deaths increases by a factor of *e*^0.33^ = 1.39 per day. There were highly statistically significant differences between upper and lower estimates (Fig. 1). Although our time series approach allowed us to estimate *r*_0_ at the start of even small epidemics, we anticipated two factors that could potentially affect our estimates of *r*_0_ that are not likely to be useful in explaining future spread rates. The first factor is the timing of the onset of county-level epidemic: 35% of the local outbreaks started after the declaration of COVID-19 as a pandemic by the WHO on 11 March, 2020^17^, and thus we anticipated estimates of *r*_0_ to decrease with the Julian date of outbreak onset. Change in human behaviors caused by public awareness about COVID-19 at the outbreak onset will not necessarily predict future rates of spread. We used the second factor, the size of the population encompassed by the time series, to factor out statistical bias from the time series analyses. Simulation studies showed that estimates for time series with low death counts were downward biased (Supplementary Fig. S2). Because for a given spread rate *r*(*t*) the total number of deaths in a time series should be proportional to the population size, we used population size as a covariate to remove bias. In addition to these two factors that we do not think have strong predictive value for the future rate of spread, we also anticipated effects of population density and spatial autocorrelation. Therefore, we regressed *r*_0_ against outbreak onset, population size and population density, and included spatially autocorrelated error terms.

**Fig. 1.**
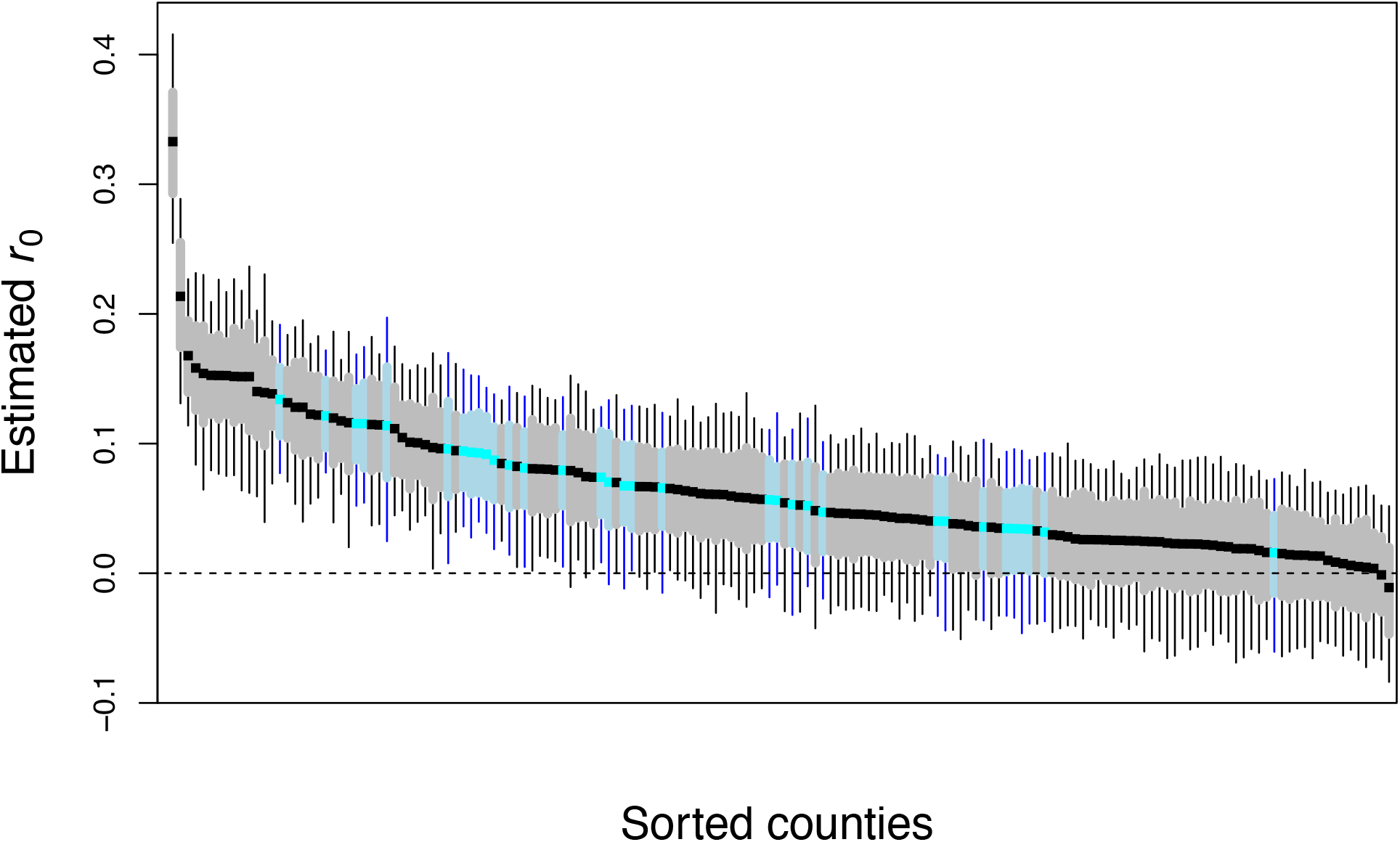
Estimates of initial spread rate, *r*_0_. The figure shows *r*_0_ point estimates (in black), sorted by magnitude, for 124 counties (gray) and 36 county-aggregates (blue), with 66% (bars) and 95% (whiskers) bootstrapped confidence intervals.

### Explaining variation in *r*_0_

The regression analysis showed highly significant effects of all four factors (Table 1), and each factor had a substantial partial R^2^_pred_^18^. The overall R^2^_pred_ was 0.70, so most of the county-to-county variance was explained. We calculated corrected *r*_0_ values, factoring out outbreak onset and population size, by standardizing the *r*_0_ values to 11 March, 2020, and to the most populous county (for which the estimates of *r*_0_ are likely best). Counties with low to medium population density never had high corrected *r*_0_ values, suggesting that population density sets an upper limit on the rate of spread of COVID-19 (Fig. 2A), in agreement with expectations and published results^1,19^. Nonetheless, despite the unequivocal statistical effect of population density (*P* < 10^−8^, Table 1), the explanatory power was not high in comparison to the entire model (partial R^2^_pred_ = 0.14), probably because population density at the scale of counties will be only roughly related to contact rates among people. The contact rates will likely depend on a wide variety of factors, such as transmission through schools, social gatherings, and nursing homes.

**Table 1.** For 160 county and county-aggregates, results of the regression of the estimates of the initial spread rate, *r*_0_, against (i) the date of outbreak onset, (ii) total population size and (iii) population density, in which (iv) spatial autocorrelation is incorporated into the residual error. Transforms of population size and density were selected to best-fit the data and satisfy linearity assumptions. The coefficient column contains the estimate of the regression parameters with their associated t-tests; spatial autocorrelation is characterized by a range and nugget for regional and local sources of variation, and their joint significance is given by a likelihood ratio test. For the overall model, R^2^_pred_ = 0.70, and the residual standard error is 1.19.

| | Coefficient | SE | t | P | partial $R^2_{\text{pred}}$ |
| --- | --- | --- | --- | --- | --- |
| **onset** | -0.0019 | 0.0004 | -4.59 | $10^{-4}$ | 0.11 |
| **log(size)** | 0.0247 | 0.0028 | 8.92 | $< 10^{-8}$ | 0.36 |
| **density<sup>1/4</sup>** | 0.025 | 0.0028 | 8.92 | $< 10^{-8}$ | 0.14 |
| **space** | range = 5.71<br>nugget = 0.33 | | $\chi^2_2 = 73$ | $< 10^{-8}$ | 0.48 |

**Fig. 2.**
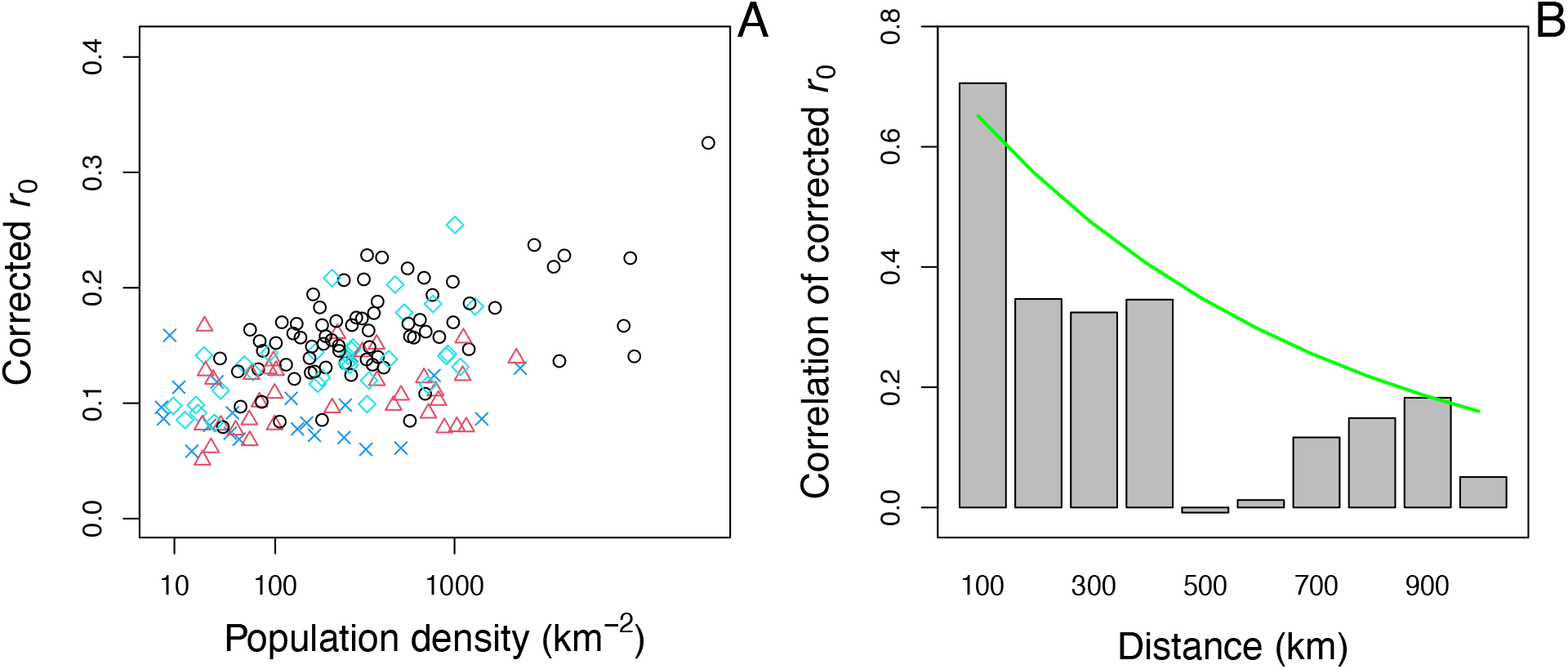
Estimates of initial spread rates, *r*_0_, after correcting for the effects of outbreak onset and population size. **(A)** Effect of population density: Northeast, black circles; Midwest, cyan diamonds; South, blue x’s; West, red triangles. **(B)** Effect of spatial proximity depicted by computing correlations in bins representing 0-100 km, 100-200 km, etc. The line gives the correlation of the residuals from the fitted regression.

Spatial autocorrelation had strong power in explaining variation in *r*_0_ among counties (partial R^2^_pred_ = 0.48, Table 1) and occurred at the scale of hundreds of kilometers (Fig. 2B). This spatial autocorrelation might reflect differences in public responses to COVID-19 across the USA not captured by the variable in the regression model for outbreak onset. For example, Seattle, WA, reported the first positive case in the USA, on 15 January, 2020, and there was a public response before deaths were recorded^20^. In contrast, the response in New York City was delayed, even though the outbreak occurred later than in Seattle^21^. Spatial autocorrelation could also be caused by movement of infected individuals. However, movement would only lead to autocorrelation in our regression analysis if many of the reported deaths were of people infected outside the county; while some deaths were likely caused by infections from outside counties, privacy restrictions on case data make these data hard to obtain, and we assume that such deaths are a small proportion of the total. A further possibility is that spatial variation in the rate of spread of COVID-19 reflects spatial variation in the occurrence of different genetic strains of SARS-CoV-2.

To investigate whether spatial autocorrelation could potentially be caused by different strains of SARS-CoV-2 differing in transmissibility, we analyzed publicly available information about genomic sequences from the GISAID metadata^22^. Scientific debate has focused on the role of the G614 mutation in the spike protein gene (D614G) to increase the rate of transmission of SARS-CoV-2^23^. We therefore asked whether the proportion of strains containing the G614 mutation was associated with higher rates of COVID-19 spread. Because the genomic samples are only located to the state level, we performed the analysis accordingly, for each state selecting the *r*_0_ from the county or county-aggregate with the highest number of deaths (and hence being most likely represented in the genomic samples). We further restricted genomic samples to those collected within 30 days following the outbreak onset we used to select the data for time-series analyses, and we required at least five genomic samples per state. This data handling resulted in 28 states available for analysis. We again used our regression model (Eq. 3), now including the proportion of strains having the G614 mutation instead of spatial location. The proportion of samples containing the G614 mutation had a positive effect on *r*_0_ (*P* = 0.016, Table S2). The low proportion of strains containing the G614 mutation in the Pacific Northwest and the Southeast were associated with lower values of *r*_0_ (Fig. 3). Before analyzing the full GISAID data, we analyzed a subset from Nextstrain^24^ naïvely, without engaging the specific hypothesis that the G614 mutation increased transmission. This naïve analysis picked strain 19B as having a lower transmission rate than other strains (*P* = 0.014, Supplementary Methods: Analysis of Nextstrain metadata of SARS-CoV-2 strains). Strain 19B does not have the G614 mutation, although strain 19A (also without the G614 mutation) did not have lower transmission than G614-containing strains, suggesting possible differences among strains separate from or in addition to the G614 mutation^25^.

**Fig. 3.**
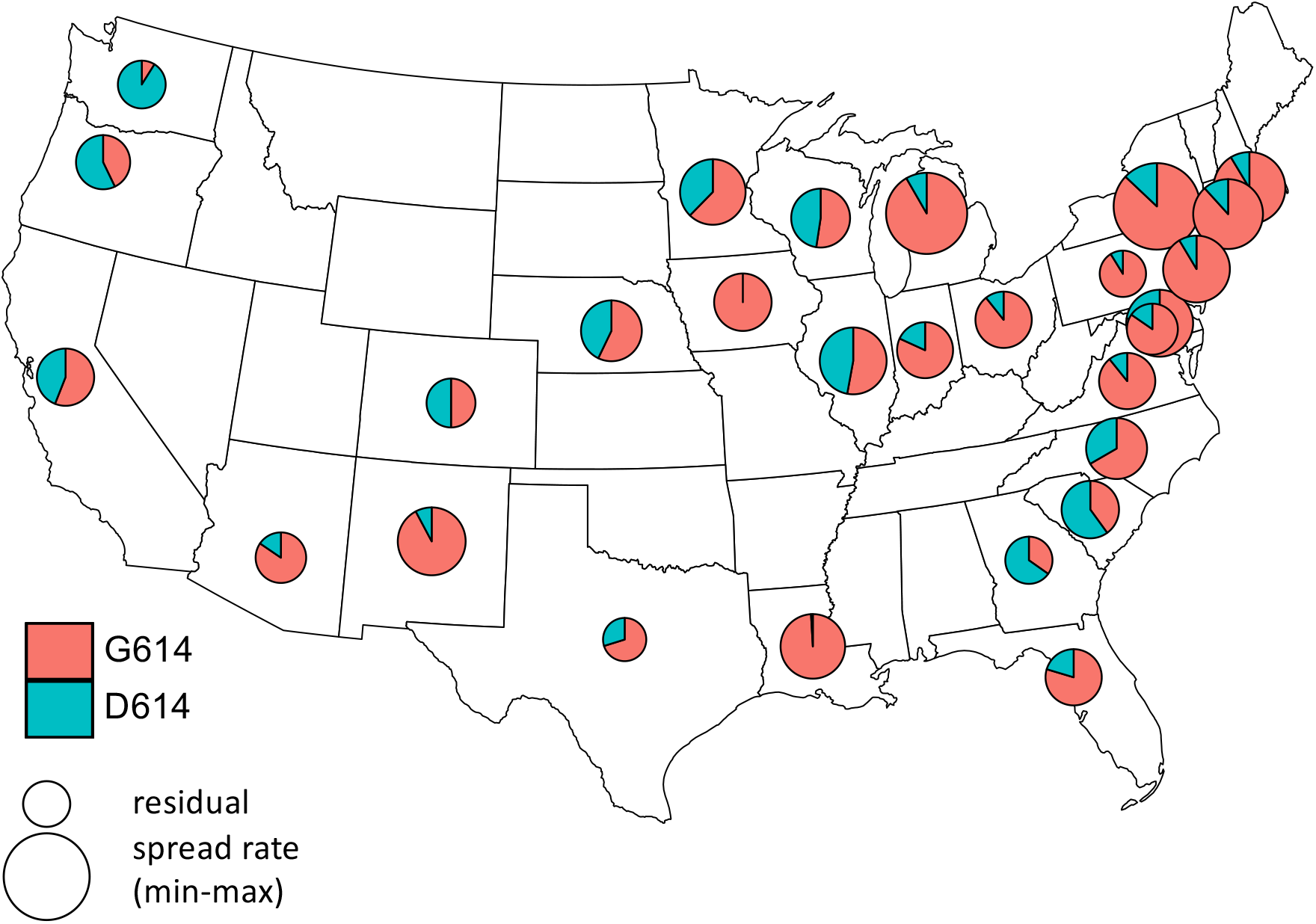
Spatial distribution of strains of SARS-CoV-2 having the G614 mutation in the spike gene at the outbreak onset among states. Pie charts give the proportion of samples in states collected within 30 days following the outbreak onset that are in the G clades (red)^22^. The size of the pie is proportional to the residual values of *r*_0_ after removing the effects of the timing of outbreak onset, population size represented by the time series, and population density. For each state, we used the estimate of *r*_0_ corresponding to the county or county-aggregate that had the greatest number of deaths.

Higher transmissibility of strains containing the G614 mutation is also suggested by its increasing prevalence in strains in the USA^26^. Nonetheless, our analyses give no information about the mechanisms explaining differences in spread rates among strains. A consensus on the potential impact of SARS-CoV-2 mutations is still lacking^23^: some studies present evidence for a differential pathogenicity and transmissibility^27,28^, while others conclude that mutations might be mostly neutral or even reduce transmissibility^29^. Our analyses call for further investigation to better understand the potential link between viral genomic variation and its impact on transmission and mortality^30^.

To check whether there are other factors that might explain variation in our estimates of *r*_0_ among counties, we investigated additional population characteristics^31-38^ that might be expected to affect the initial spread rate of COVID-19: (i) proportion of the population over 65, (ii) adult obesity, (iii) diabetes, (iv) education, (v) income, (vi) poverty, (vii) economic equality, (viii) race, and (ix) political leaning (Table S4). The first three characteristics likely affect morbidity^39^, although it is not clear how higher morbidity could affect the spread rate. The remaining characteristics might affect health outcomes and responses to public health interventions; for example, education, income, and poverty might all affect the need for individuals to work in jobs that expose them to greater risks of infection. Nonetheless, because we focused on the early spread of COVID-19, we anticipated that these characteristics would have minimal effects. Despite the potential for all nine characteristics to affect estimates of *r*_0_, we found that none was a statistically significant predictor of *r*_0_ when taking the four main factors into account (all *P* > 0.1). We also repeated all of the analyses on estimates of *r*(*t*) after COVID-19 was broadly established in the USA (5 May, 2020, assuming an average time between infection and death of 18 days) (Table S5). The corresponding R^2^_pred_ = 0.40, largely driven by a large positive effect of the date of outbreak onset. The absence of significant effects of the additional population characteristics on *r*_0_, and the lower explanatory power of the model on *r*(*t*) at the end of the time series, underscore the importance of population density and spatial autocorrelation in predicting county-level values of *r*_0_.

### Extrapolating R_0_ to all counties

In the regression model (Table 1), the standard deviation of the residuals was 1.18 times higher than the average standard error of the estimates of *r*_0_. This implies that the uncertainty of an estimate of *r*_0_ from the regression is only slightly higher than the uncertainty in the estimate of *r*_0_ from the time series itself; the fixed terms (ignoring spatial autocorrelation) in the regression model explain 71% (= 1/1.19^2^) of the explainable variance in *r*_0_. Therefore, using estimates from death count time series from other counties will give estimates of *r*_0_ for a focal county (lacking reliable estimates) that are almost as precise as the estimate from the county’s time series. We used the regression to extrapolate values of R_0_, for all 3109 counties in the conterminous USA (Fig. 4, Table S1). The high predictability of *r*_0_, and hence R_0_, from the regression is seen in the comparison between R_0_ calculated from the raw estimates of *r*_0_ (Fig. 4A) and R_0_ calculated from the corrected *r*_0_ values (Fig. 4B). Extrapolation from the regression model makes it possible not only to get refined estimates for the counties that were aggregated in the time-series analyses; it also gives estimates for counties within states with so few deaths that county-aggregates could not be analyzed (Fig. 4C,D). The end product is a map of estimated R_0_ values for the conterminous USA (Fig. 4E).

**Fig. 4.**
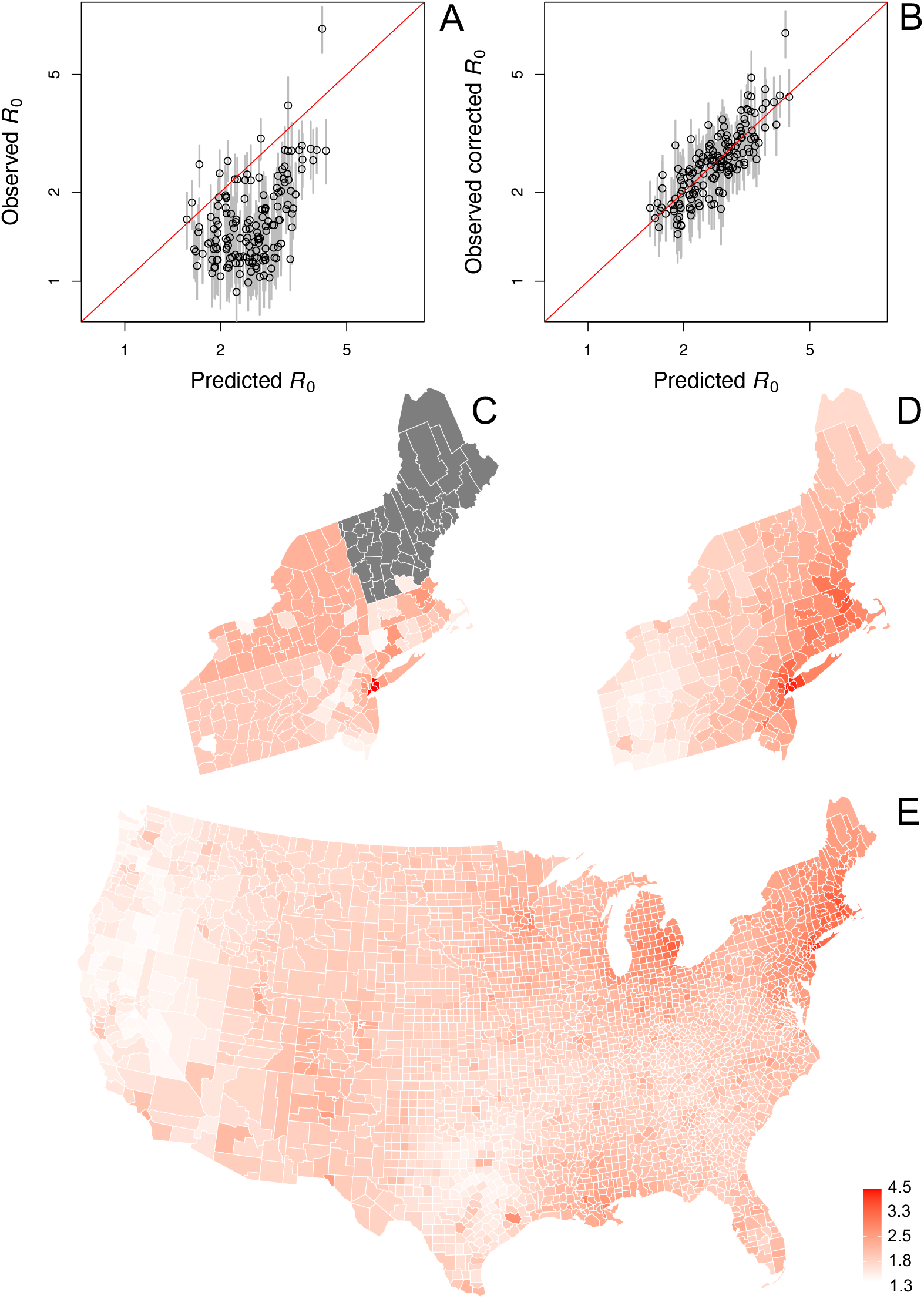
Prediction of R_0_ values for all 3109 counties in the conterminous USA. **(A**,**B)** Raw and corrected estimates of R_0_ for 160 counties and county-aggregates. The predicted R_0_ values are obtained from the regression model, with corrections to standardize values to an outbreak onset of 11 March, 2020, and a population size equal to the most populous county. Comparing the raw estimates of R_0_ (A) and the corrected R_0_ values (B) shows the predictive power of the regression analysis. We thus used the regression model to predict R_0_ for all counties. **(C**,**D)** To illustrate the prediction process for the northeastern states, the raw estimates (C) are all the same for county-aggregates and could not be made for some states (gray). In contrast, the predictability R_0_ in the regression model allows for better estimates (D). **(E)** This makes it possible to extend estimates of R_0_ to all 3109 counties in the conterminous USA.

## Discussion

It is widely understood that different states and counties in the USA, and different countries in the world, have experienced COVID-19 epidemics differently. Our analyses have put numbers on these differences in the USA. The large differences argue for public health interventions to be designed at the county level. For example, the vaccination coverage in the most densely populated area, New York City, needed to prevent future outbreaks of COVID-19 will be much greater than for sparsely populated counties. Therefore, once vaccines are developed, they should be distributed first to counties with high R_0_. Similarly, if vaccines are not developed quickly and non-pharmaceutical public health interventions have to be re-instated during resurgent outbreaks, then counties with higher R_0_ values will require stronger interventions. As a final example, county-level R_0_ values can be used to assess the practicality of contact-tracing of infections, which become impractical when R_0_ is high^40,41^.

Estimating county-level values of R_0_ at the start of the epidemic faces statistical challenges that our analyses have tried to address. We used death counts, rather than cases reported from testing, because particularly at the start of the epidemic, testing was limited and heterogeneous among states and counties. Nonetheless, death counts are not perfect, because different criteria could be used by different counties to ascribe deaths to SARS-CoV-2. Furthermore, evidence suggests that “excess deaths” have occurred in comparison to historical data^12^ and that these excess deaths are likely due to the mis-attribution to causes other than SARS-CoV-2. Nonetheless, we estimated R_0_ from the spread rate of the disease (equation 1), which depends on the change in the number of recorded deaths from one day to the next. This change in death counts should be insensitive to the criteria used to ascribe death to SARS-CoV-2, and although there are undoubtedly mistakes and omissions, our statistical methods account for this measurement error.

We present our county-level estimates of R_0_ as preliminary guides for policy planning, while recognizing the myriad other epidemiological factors (such as mobility^42-44^) and political factors (such as legal jurisdictions^45^) that must shape public health decisions^3,46-48^. Although we have emphasized the high predictability of R_0_ among counties in the USA, our estimates of R_0_ will be under-estimates for some regions if there are changes in the transmissibility of strains (Fig. 3). This uncertainty underscores the need for more information about strain differences affecting SARS-CoV-2 transmission^23,25^.

We recognize the importance of following the day-to-day changes in death and case rates, and short-term projections used to anticipate hospital needs and modify public policies^49-51^. Looking back to the initial spread rates, however, gives a window into the future and what public health policies will be needed when COVID-19 is endemic.

## Methods

### 1. Data selection and handling

#### 1.1 Death data

For mortality due to COVID-19, we used time series provided by the New York Times^11^. We selected the New York Times dataset because it is rigorously curated. We analyzed separately only counties that had records of 100 or more deaths. The threshold of 100 was a balance between including more counties and obtaining reliable estimates of *r*(*t*). Preliminary simulations showed that time series with low numbers of deaths would bias *r*(*t*) estimates (Supplementary Fig. S2). However, we did not want to use the maximum number of deaths as a selection criterion, because this could lead to selection of counties based on data from a single day. It would also involve some circularity, because the information obtained, *r*(*t*), would be directly related to the criterion used to select data sets. Therefore, we used the threshold of 100 cumulative deaths. The District of Columbia was treated as a county. Also, because the New York Times dataset aggregated the five boroughs of New York City, we treated them as a single county. For counties with fewer than 100 deaths, we aggregated mortality to the state level to create a single time series. For thirteen states (AK, DE, HI, ID, ME, MT, ND, NH, SD, UT, VM, WV, and WY), the aggregated time series did not contain 100 or more deaths and were therefore not analyzed.

#### 1.2 Explanatory county-level variables

County-level variables were collected from several public data sources^32-38^. We selected socio-economic variables *a priori* in part to represent a broad set of population characteristics.

### 2. Time series analysis

#### 2.1 Time series model

We used a time-varying autoregressive model^13,52,53^ designed explicitly to estimate the rate of increase of a variable using nonlinear, state-dependent error terms. We assume in our analyses that the susceptible proportion of the population represented by a time series is close to one, and therefore there is no decrease in the infection rate caused by a pool of individuals who were infected, recovered, and were then immune to further infection.

The model is

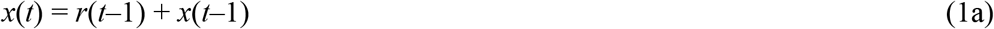

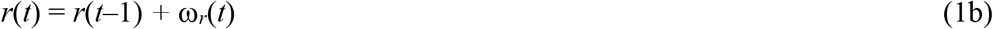

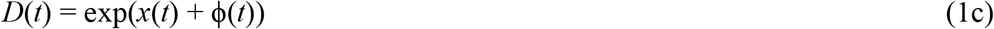

Here, *x*(*t*) is the unobserved, log-transformed value of daily deaths at time *t*, and *D*(*t*) is the observed count that depends on the observation uncertainty described by the random variable ϕ(*t*). Because a few of the datasets that we analyzed had zeros, we replaced zeros with 0.5 before log-transformation. The model assumes that the death count increases exponentially at rate *r*(*t*), where the latent state variable *r*(*t*) changes through time as a random walk with ω_*r*_(*t*) ∼ N(0, σ^2^r). We assume that the count data follow a quasi-Poisson distribution. Thus, the expectation of counts at time *t* is exp(*x*(*t*)), and the variance is proportional to this expectation.

We fit the model using the Kalman filter to compute the maximum likelihood^54,55^. In addition to the parameters σ^2^r and σ^2^_ϕ_, we estimated the initial value of *r*(*t*) at the start of the time series, *r*_0_, and the initial value of *x*(*t*), *x*_0_. The estimation also requires an assumption for the variance in *x*_0_ and *r*_0_, which we assumed were zero and σ^2^*r*, respectively. In the validation using simulated data (Supplementary Methods: Simulation model), we found that the estimation process tended to absorb σ^2^*r* to zero too often. To eliminate this absorption to zero, we imposed a minimum of 0.02 on σ^2^*r*, which eliminated the problem in the simulations.

#### 2.2 Parametric bootstrapping

To generate approximate confidence intervals for the time-varying estimates of *r*(*t*) (Eq. 1b), we used a parametric bootstrap designed to simulate datasets with the same characteristics as the real data that are then refit using the autoregressive model. We used bootstrapping to obtain confidence intervals, because an initial simulation study showed that standard methods, such as obtaining the variance of *r*(*t*) from the Kalman filter, were too conservative (the confidence intervals too narrow) when the number of counts was small. Furthermore, parametric bootstrapping can reveal bias and other features of a model, such as the lags we found during model fitting (Supplementary Fig. S1A,B).

Changes in *r*(*t*) consist of unbiased day-to-day variation and the biased deviations that lead to longer-term changes in *r*(*t*). The bootstrap treats the day-to-day variation as a random variable while preserving the biased deviations that generate longer-term changes in *r*(*t*). Specifically, the bootstrap was performed by calculating the differences between successive estimates of *r*(*t*), Δ*r*(*t*) = *r*(*t*) – *r*(*t*-1), and then standardizing to remove the bias, Δ*r*_*s*_(*t*) = Δ*r*(*t*) – E[Δ*r*(*t*)], where E[] denotes the expected value. The sequence Δ*r*_*s*_(*t*) was fit using an autoregressive time-series model with time lag 1, AR(1), to preserve any shorter-term autocorrelation in the data. For the bootstrap a new time series was simulated from this AR(1) model, Δ*ρ*(*t*), and then standardized, Δ*ρ*_*s*_(*t*) = Δ*ρ*(*t*) – E[Δ*ρ*(*t*)]. The simulated time series for the spread rate was constructed as *ρ*(*t*) = *r*(*t*) + Δ*ρ*_*s*_(*t*)/ 2^1/2^, where dividing by 2^1/2^ accounts for the fact that Δ*ρ*_*s*_(*t*) was calculated from the difference between successive values of *r*(*t*). A new time series of count data, *ξ*(*t*), was then generated using equation 1 with the parameters from fitting the data. Finally, the statistical model was fit to the reconstructed *ξ*(*t*). In this refitting, we fixed the variance in *r*(*t*), σ^2^*r*, to the same value as estimated from the data. Therefore, the bootstrap confidence intervals are conditional of the estimate of σ^2^*r*.

#### 2.3. Calculating R_0_

We derived estimates of *R*(*t*) directly from *r*(*t*) using the Dublin-Lotka equation^16^ from demography. This equation is derived from a convolution of the distribution of births under the assumption of exponential population growth. In our case, the “birth” of COVID-19 is the secondary infection of susceptible hosts leading to death, and the assumption of exponential population growth is equivalent to assuming that the initial rate of spread of the disease is exponential, as is the case in equation 1. Thus,

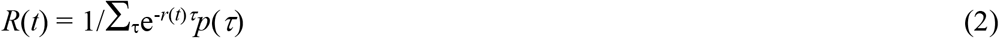

where *p*(τ) is the distribution of the proportion of secondary infections caused by a primary infection that occurred τ days previously. We used the distribution of *p*(τ) from Li et al.^56^ that had an average serial interval of T_0_ = 7.5 days; smaller or larger values of T_0_, and greater or lesser variance in *p*(τ), will decrease or increase *R*(*t*) but will not change the pattern in *R*(*t*) through time. Note that the uncertainty in the distribution of serial times for COVID-19 is a major reason why we focus on estimating *r*_0_, rather than R_0_: the estimates of *r*_0_ are not contingent on time distributions that are poorly known. Computing *R*(*t*) from *r*(*t*) also does not depend on the mean or variance in time between secondary infection and death. We report values of *R*(*t*) at dates that are offset by 18 days, the average length of time between initial infection and death given by Zhou et al.^57^.

#### 2.4. Initial date of the time series

Many time series consisted of initial periods containing zeros that were uninformative. As the initial date for the time series, we chose the day on which the estimated daily death count exceeded 1. To estimate the daily death count, we fit a Generalized Additive Mixed Model (GAMM) to the death data while accounting for autocorrelation and greater measurement error at low counts using the R package mgcv^58^. We used this procedure, rather than using a threshold of the raw death count, because the raw death count will include variability due to sampling small numbers of deaths. Applying the GAMM to “smooth” over the variation in count data gives a well-justified method for standardizing the initial dates for each time series.

#### 2.5. Validation

We performed extensive simulations to validate the time-series analysis approach (Supplementary Methods: Simulation model).

### 3. Regression analysis for *r*_0_

We applied a Generalized Least Squares (GLS) regression model to explain the variation in estimates of *r*_0_ from the 160 county and county-aggregate time series:

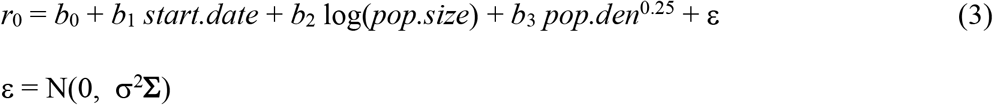

where *start*.*date* is the Julian date of the start of the time series, log(*pop*.*size*) and *pop*.*den*^0.25^ are the log-transformed population size and 0.25 power-transformed population density of the county or county-aggregate, respectively, and ε is a Gaussian random variable with covariance matrix σ^2^**Σ**. We used the transforms log(*pop*.*size*) and *pop*.*den*^0.25^ to account for nonlinear relationships with *r*_0_, and we selected these transforms to give the highest maximum likelihood of the overall regression. The covariance matrix contains a spatial correlation matrix of the form **C** = *u***I** + (1-*u*)**S**(*g*) where *u* is the nugget and **S**(*g*) contains elements exp(*-d*_*ij*_*/g*), where *d*_*ij*_ is the distance between spatial locations and *g* is the range^59^. To incorporate differences in the precision of the estimates of *r*_0_ among time series, we weighted by the vector of their standard errors, **s**, so that **Σ** = diag(**s**) * **C** * diag(**s**), where * denotes matrix multiplication. With this weighting, the overall scaling term for the variance, σ^2^, will equal 1 if the residual variance of the regression model matches the square of the standard errors of the estimates of *r*_0_ from the time series. We fit the regression model with the function gls() in the R package nlme^60^.

To make predictions for new values of *r*_0_, we used the relationship

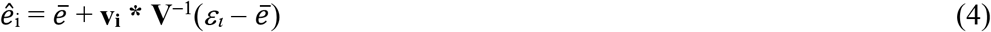

where ε_*l*_ is the GLS residual for data *i, ê*_i_ is the predicted residual, *ē* is the mean of the GLS residuals, **V** is the covariance matrix for data other than *i*, and **v**_**i**_ is a row vector containing the covariances between data *i* and the other data in the dataset^61^. This equation was used for three purposes. First, we used it to compute R^2^_pred_ for the regression model by removing each data point, recomputing *ê*_i_, and using these values to compute the predicted residual variance^18^. Second, we used it to obtain predicted values of *r*_0_, and subsequently R_0_, for the 160 counties and county-aggregates for which *r*_0_ was also from time series. Third, we used equation (4) to obtain predicted values of *r*_0_, and hence predicted R_0_, for all other counties. We also calculated the variance of the estimates from^61^

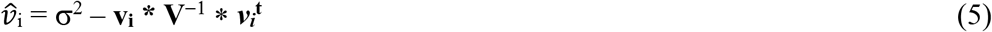

Predicted values of R_0_ were mapped using the R package usmap^62^.

### 4. Regression analysis for SARS-CoV-2 effects on *r*_0_

The GISAID metadata^22^ for SARS-CoV-2 contains the clade and state-level location for strains in the USA; strains G, GH, and GR contain the G614 mutation. For each state, we limited the SARS-CoV-2 genomes to those collected no more than 30 days following the onset of outbreak that we used as the starting point for the time series from which we estimated *r*_0_; from these genomes (totaling 5290 from all states), we calculated the proportion that had the G614 mutation. Only twenty-eight states had five or more genomes, so we limited the analyses to these states. For each state, we selected the estimates of *r*_0_ from the county or county-aggregate representing the greatest number of deaths. We fit these estimates of *r*_0_ with the weighted Least Squares (LS) model as in equation (3) with additional variables for strain. Figure 3 was constructed using the R packages usmap^62^ and scatterpie^63^.

### 5. Statistics and Reproducibility

The statistics for this study are summarized in the preceding sections of the Methods. No experiments were conducted, so experimental reproducibility is not an issue. Nonetheless, we repeated analyses using alternative datasets giving county-level characteristics, and also an alternative dataset on SARS-CoV-2 strains (Supplementary Methods: Analysis of Nextstrain metadata of SARS-CoV-2 strains), and all of the conclusions were the same.

## Supporting information

Source code and compiled data

## Data Availability

Data and code have been uploaded with an earlier version of this manuscript.

## Acknowledgments

We thank Steve R. Carpenter, Volker C. Radeloff, and Monica M. Turner for comments on the manuscript.

## Author contributions

A.R.I and C.B. designed the study, and A.R.I. led the analyses and writing of the manuscript.

## Data availability

All data are included in the text by referencing to the original sources.

## Code availability

Data and R code for the analyses are presented in the online Supplementary Information.

## Supplementary Information

### Supplementary Methods

#### Overview of Statistical Methods

The rate of spread of a disease in a population at the early phase of an epidemic, *r*_0_, when the entire population is susceptible depends on the basic reproduction number, R_0_, giving the number of secondary infections produced per infected individual, and the distribution of the time between primary and secondary infections. Thus, if the spread rate and distribution of infection times can be estimated, R_0_ can then be calculated. Our strategy is to estimate *r*_0_ as the most direct parameter associated with the dynamics of an epidemic, and then subsequently estimate R_0_. The advantages of calculating *r*_0_ include: (i) it captures all of the real-life complexities that affect R_0_ by simply observing what happened in real life, and (ii) it uses data that are (tragically) becoming more prevalent. The challenges include (i) the changes in *r*(*t*) that are to be expected (and hoped for) as people and governments respond to lessen the spread, and (ii) the statistical challenges and uncertainties of determining rates of disease spread when the numbers of deaths are still low.

We developed and tested statistical methods to overcome the two challenges of estimating R_0_ from death data. Because the rate of spread of a disease may change rapidly in response to actions that are taken to reduce disease transmission, we used a time-varying autoregressive model that allows for the rate of spread to change through time, *r*(*t*). Other models take a related approach^1,2^. The second challenge is that the counts of deaths at the beginning of an epidemic are low. To account for this, the time-series model includes increased uncertainty (measurement error) that depends on the time-varying estimate of the number of deaths. Standard (asymptotic) approaches often have poor statistical properties (type I errors, correctly calculated confidence intervals) when sample sizes are small^3^. Therefore, we use bootstrapping^4^ in which simulation time series are reconstructed to share the same pattern as the observed time series; a large number of simulated time series are then fit using the same statistical model as used to fit the original data. This bootstrapping procedure thus gives estimates and confidence intervals for model fit to the real data. Note that our approach is frequentist, in comparison to the majority of models that use a Bayesian framework.

Our approach focuses on estimating the time-varying rate of spread, *r*(*t*), of the number of deaths. Our rationale is that, for statistical fitting, it is better to keep the model as simple as possible, rather than “building in” assumptions about the processes of infection, reporting, and death. Our simple phenomenological model uses the same data as more-complicated, process-based models, and therefore both approaches ultimately rely on the same information. The simpler approach, however, does not depend on assumptions about the infection processes. Instead, after estimating *r*_0_, we computed R_0_ as 1/Σ_τ_e^-*r*(*t*)τ^*p*(τ), where τ is the number days after initial infection, and *p*(τ) is the proportion of secondary infections produced per infected individual at τ _5_. This expression assumes that deaths (removal of individuals from the population) occur after all secondary infections have occurred. We used the distribution of *p*(τ) that was estimated using contact tracing in Wuhan, China^6^.

To validate the statistical method, we constructed a simulation model of the transmission process and spread of infections iterated on a daily time scale. Our simulations considered scenarios in which the transmission rate changed through time either in steps or gradually to capture the extremes of possible changes in real *R*(*t*). We varied the initial R_0_ and duration of simulations to produce epidemics that qualitatively match the county data we analyzed. Changes in our estimates of *r*(*t*) tended to lag behind changes in the true (simulated) value of *r*(*t*) (gray line and regions in Supplementary Fig. S1A,B), and therefore we also estimated *r*(*t*) in the reverse direction (blue line and regions in Supplementary Fig. S1A,B). For the estimate of the initial *r*_0_, we averaged the estimates from the forward and reverse time series. For the scenario of step changes in *R*(*t*) (Supplementary Fig. S1C), the estimates were unbiased and had accurate confidence intervals, although for the scenario of gradual changes (Supplementary Fig. S1D), there was some downwards bias. Nonetheless, the estimates of initial R_0_ captured the order of simulations according to the true R_0_. In contrast, fitting the same time series with a commonly used Bayesian model that incorporates the transmission process given in the R package EpiEstim^7^ gave estimates that poorly reflect the true (simulated) initial R_0_ (Supplementary Fig. S1E,F).

We also used the simulation model to investigate the properties of the statistical method when the number of deaths was low, as occurred in some time series. Reducing the simulated values of R_0_ reveals that the estimates of *r*_0_ become biased downwards when the maximum number of reported deaths per day drops below 15 (Supplementary Fig. S2A). This is due to the time series containing too little information about the rate of increase in the number of mortalities for accurate estimates. Because we did not think that our method (or any other) could overcome this challenge, we incorporated population size encompassed by a time series in the subsequent regression analysis. We used population size rather than the maximum number of deaths, because this would introduce a confounding effect: time series with higher *r*_0_ will likely have higher numbers of deaths.

In order to extrapolate the estimates of R_0_ from 160 time series to the remaining counties in the conterminous USA, we *a priori* selected four predictors. We selected population size encompassed by the time series to account for possible downwards bias in sparse datasets. We selected the Julian date of the outbreak onset to factor out public and private responses to COVID-19. We included population density, because it could potentially affect transmission rates. Population size and density were weakly and negatively correlated among the 160 time series (Pearson correlation between log population size and log density = –0.25), and therefore there were no problems with multicollinearity. Finally, the regression model included spatial autocorrelation based on the latitude and longitude of the population-weighted midpoint of the counties or county aggregates. Because the regression model had residual variance that was only slightly higher than the variance of the estimates of *r*_0_ that the regression predicted, the precision of the estimates from the regression for the counties without time series will be on par with the precision of the counties with time series.

#### Simulation model

To assess the robustness of the statistical model, we built a simulation model of a hypothetical epidemic. The simulation model tracks the epidemic on a daily time scale and explicitly includes the time period from infection to subsequent transmission (infectiousness), and from infection to death; therefore, it is akin to a SEIR model. The simulation model was not the same as the statistical model, so the goal was to determine whether the phenomenological statistical model was capable of capturing the rate of infection spread in the process-based simulations.

The simulation model tracks the number of infected individuals on day *t* who were infected τ days previously, *X*(*t*;τ). After 25 days, they are all assumed to be recovered or dead. The probability distribution of the day on which a susceptible is infected, *p*(*t*), is given by a Weibull distribution with mean 7.5 days and standard deviation 3.4^6^ (Supplementary Fig. S3A). For an individual who dies, the day of death, *d*(*t*), is given by a Weibull distribution with mean 18.5 days and standard deviation 3.4^6^ (Supplementary Fig. S3B). Finally, for case data we need to know the time between initial infection and diagnosis, *h*(*t*), which we assume is lognormally distributed with mean 5.5 days and standard deviation 2.2^8^ (Supplementary Fig. S3C).

On day *t*, the number of new infections produced by individuals who were infected τ days earlier is *b*(*t*) *p*(τ). The term *b*(*t*) is closely related to *R*(*t*), the number of secondary infections caused per infection. However, because we allow *b*(*t*) to fluctuate on a daily basis, here we use a notation that differs from *R*(*t*). Note, however, that on average *R*(*t*) = Στ *b*(*t* + τ) *p*(τ). The total number of new infections on day *t* is given by a lognormal Poisson distribution in which the mean of the Poisson process is *b*(*t*) α(*t*) Στ *p*(τ)*X*(*t*;τ), where the lognormal random variable α(*t*) is included to represent environmental variation.

Deaths occur according to a binomial distribution for each infection age category *X*(*t*;τ), so that the probability of death of individuals that had been infected τ days earlier is (1 – *s*) β(*t*) *d*(τ), where *s* is the overall survival probability and β(*t*) is a lognormal distribution. We assume that the overall survival probability for COVID-19 is 98%; changes in this assumption had little effect on the simulation study. Once an individual dies, they are removed from the pool of individuals.

To illustrate the simulations, we assumed that the expectation of the infection rate, *b*(*t*), changes as a step function (Supplementary Fig. S4A, black line), while there is also daily variation around this expectation (Supplementary Fig. S4A, points). We also calculated *R*(*t*) from the asymptotic rate of disease spread (Supplementary Fig. S4A, red line). This shows that the expected daily infection rate, *b*(*t*), is closely related to the population-level *R*(*t*). Over the simulated time series of 60 days, we then recorded the number of deaths (Supplementary Fig. S4B) and diagnosed cases (Supplementary Fig. S4C). We initiated the simulation with a single cohort of individuals, all infected on day 1 (Supplementary Fig. S4C, filled black dot). This gives the “worst-case” situation in which the distribution of time-since-infection is far from the stable age distribution.

We fit this simulated dataset using the same procedure as we used for the real data, including the same rules to determine which day to initiate the fitted time series (Supplementary Fig. S1A). We performed a similar exercise while assuming that the expectation of the infection rate, *b*(*t*) changes geometrically, producing a linear change in *r*(*t*) (Supplementary Fig. S1B). In this particular example, the estimated values of *r*(*t*) are below the true values in the simulation in the first part of the time series. Because there was a lag in response of the estimates of *r*(*t*) relative to *b*(*t*), we fit the time series in both the forward and reversed directions, and we averaged these values (and their confidence intervals) for the final estimates. Note that this is possible in our approach, because we estimate *r*(*t*) rather than *R*(*t*).

We performed 100 simulations with the expectation of *b*(*t*) changing as either a step function (Supplementary Fig. S1C) or geometrically (Supplementary Fig. S1D), to assess the overall robustness of the modeling approach. Simulations were performed by changing the initial value of *b*(*t*). Because higher values of *b*(*t*) led to much higher numbers of deaths, we shorted the intervals between step changes and increased the decline in geometric changes in *b*(*t*) to roughly match the observed time series. Specifically, the simulated time series ranged in length from 55 to 150 days: for the case of step changes, the time series were broken into three equal periods, and for the case of geometric changes, the ending value of *b*(*t*) was kept the same. We also estimated *R*(*t*) using the R package EpiEstim under default control parameters^7^. EpiEstim has the same general structure of many of the Bayesian models that estimate *R*(*t*) directly using information about the transmission process (Supplementary Fig. S1E,F). Even though EpiEstim is structurally more complicated than our model, it tended to give values of R_0_ that were biased upwards when the true value was low, and biased downward when the true value was high. Finally, we investigated the bias in our estimates of *r*_0_ when the maximum number of deaths in a time series was low by simulating time series for 20 to 70 days, using an initial value of *b*(*t*) to correspond to R_0_ = 4, and changing the timing of step changes or the rate of geometric decline of *b*(*t*) to correspond to the length of the time series. The simulations show that the estimates of *r*_0_ are downward biased when the total numbers of counts are low (Supplementary Fig. S2).

#### Analysis of Nextstrain metadata of SARS-CoV-2 strains

In the analyses presented in the main text, we used the GISAID metadata to test the specific assumption that the G614 mutation increases the rate of spread of SARS-CoV-2. Prior to this analysis, however, we analyzed a subset of the genomic data available from Nextstrain^9^. We present this analysis here, because it was a naïve analysis that did not have a specific hypothesis about what strains might lead to higher spread rates. Instead, we asked whether the proportion of different Nextstrain clades (19A, 19B, 20A, 20B, 20C in the USA) within a population were related to *r*_0_ estimates. We used the same statistical approach as we present for the GISAID metadata, except we included the proportion of strains from clades 19A, 19B, 20A, and 20B instead of the proportion in the G clades containing mutation G614; we excluded the largest clade, 20C, because the sums of the proportions must add to one, and therefore all of the information about the distribution of strain 20C among states is contained in the distribution of the other clades. We found that the proportion of samples within clade 19B had a negative effect on *r*_0_ (*P* = 0.019, Table S3). The high proportion of strains from 19B in the Pacific Northwest and the Southeast were associated with lower values of *r*_0_ (Supplementary Fig. S5). Strain 19A, however, also does not contain the G614 mutation, and it did not have a negative effect on *r*_0_.

## Supplementary Figures and Tables

**Fig. S1.**
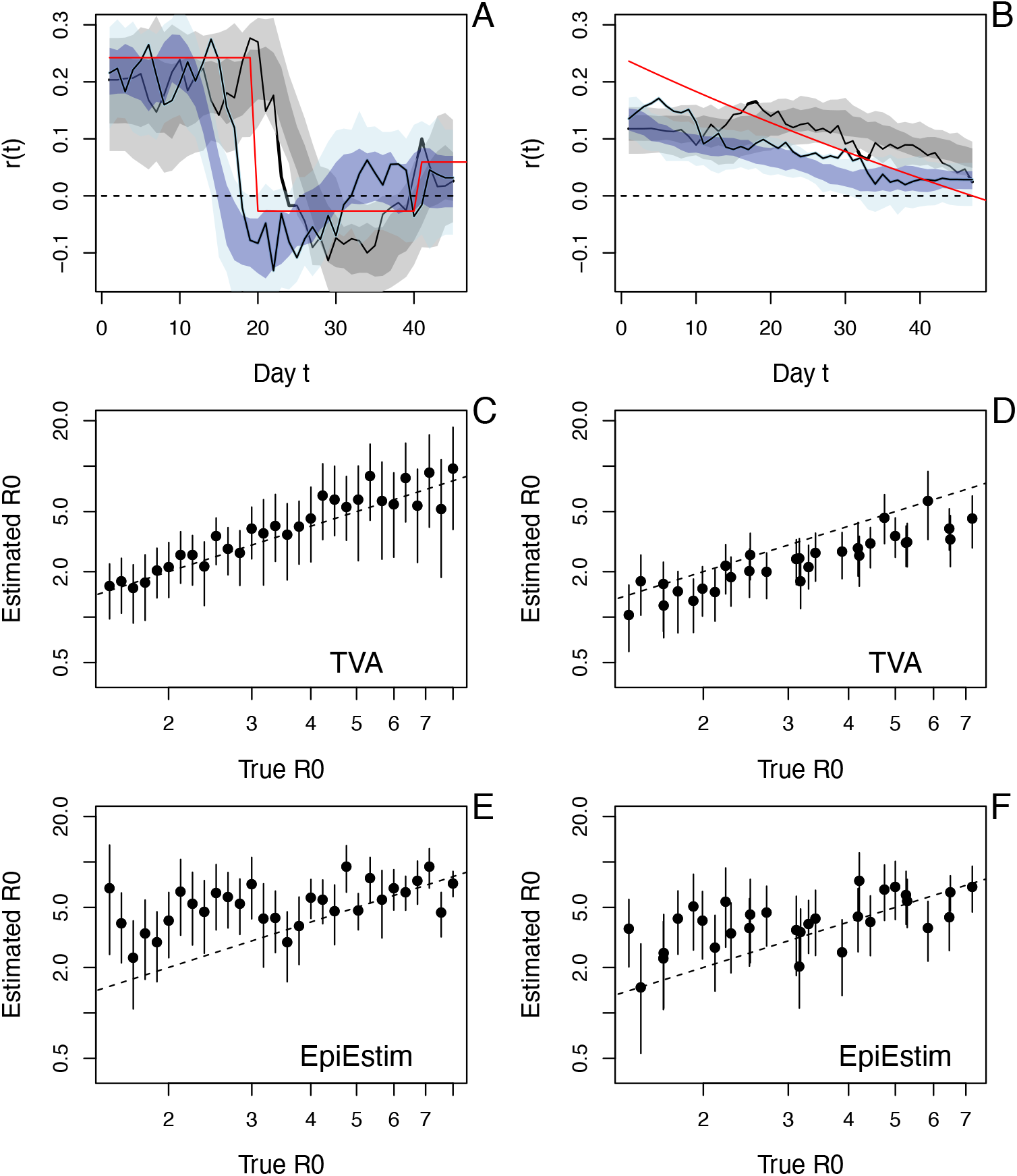
Simulation study of fitting methods to epidemic death data. Simulations were fit with the time-varying autoregression model (TVA) in the forward (black line with dark and light gray regions giving 66% and 95% approximate confidence intervals) and reverse (blue line and regions – the light blue regions are sometimes obscured) directions when the true value of R(*t*) (red line) shows either **(A)** a step or **(B)** gradual changes. For each simulation, the forward and reverse estimates were averaged to give an estimate of R_0_ with 95% confidence intervals, which are plotted against the true values of R_0_ for step **(C)** and gradual **(D)** changes in R(*t*). The same simulations with fit using EpiEstim **(E**,**F)**.

**Fig. S2.**
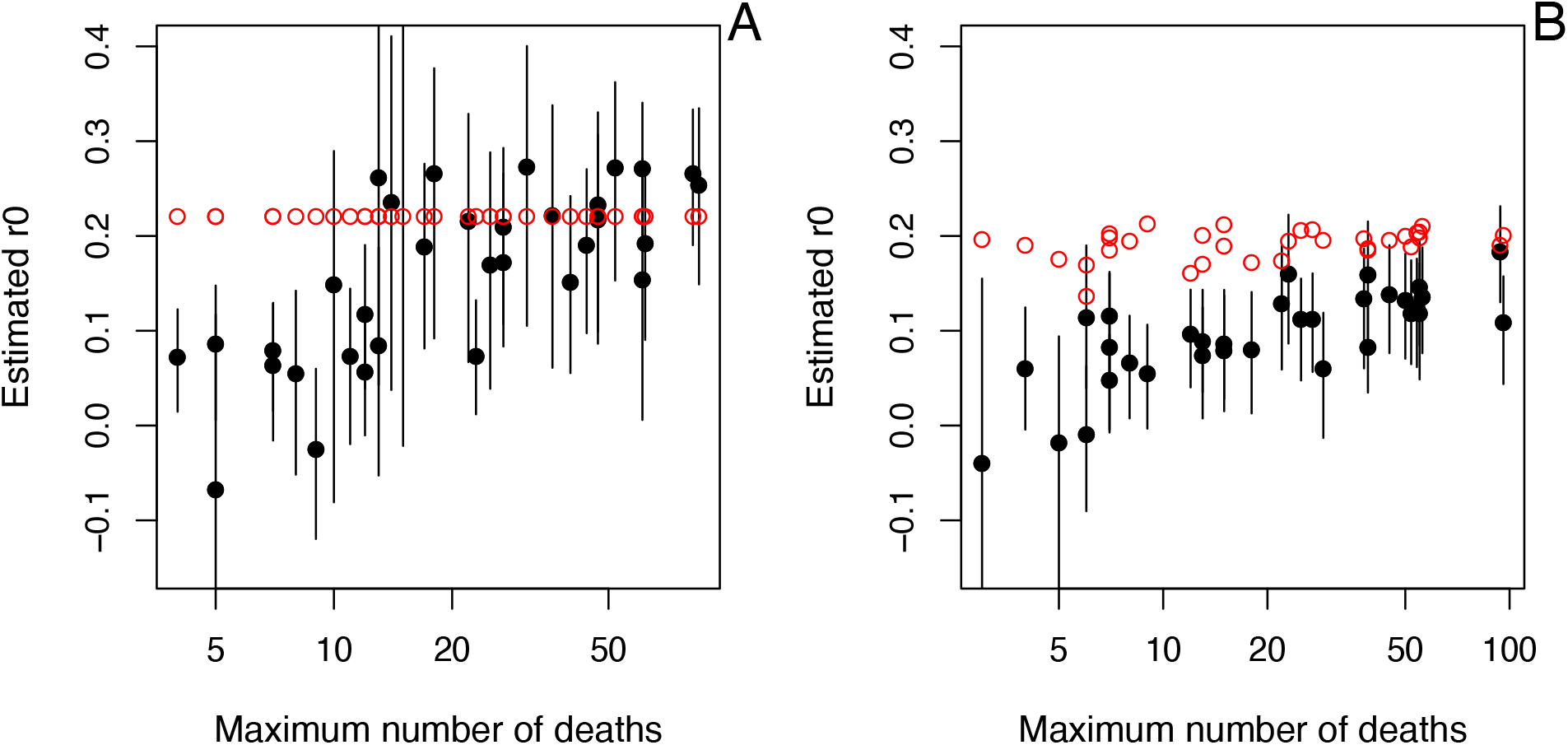
Simulation study of the estimation of *r*_0_ from the forward and reverse time-varying autoregressive model for different population sizes. Simulations following those used for Fig. S1 were performed assuming *r*(*t*) changed either **(A)** in steps or **(B)** gradually. The simulations were performed using the same initial value of *r*_0_, but the length of time of the simulation was varied to change the maximum number of deaths that occurred. Due to the stochastic nature of the simulations, the realized value of *r*_0_ when the analysis was started differed among time series when *r*(*t*) changed gradually (red points in B), while they were all 0.22 when *r*(*t*) was changed in steps (A). The median in the maximum number of deaths among the real county time series was 21.

**Fig. S3.**
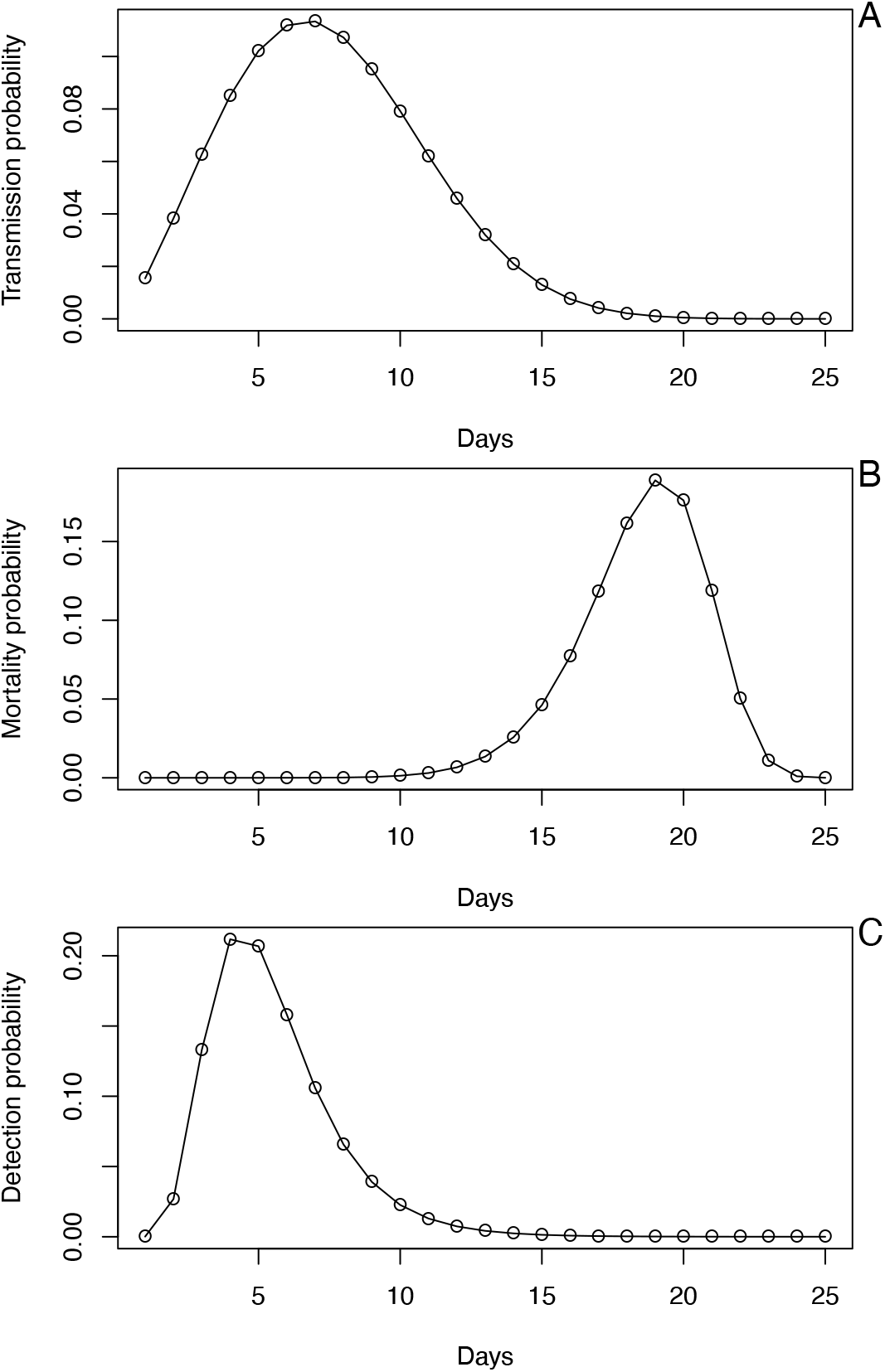
Probability distributions used in the process-based simulation model used to test methods for robustness. **(A)** The probability distribution of the day on which a susceptible is infected, *p*(*t*), given by a Weibull distribution with mean 7.5 days and standard deviation 3.4. **(B)** For an individual who dies, the day of death, *d*(*t*), which is given by a Weibull distribution with mean 18.5 days and standard deviation 3.4. **(C)** For case data, the time between initial infection and diagnosis, *h*(*t*), which is lognormally distributed with mean 5.5 days and standard deviation 2.2.

**Fig. S4.**
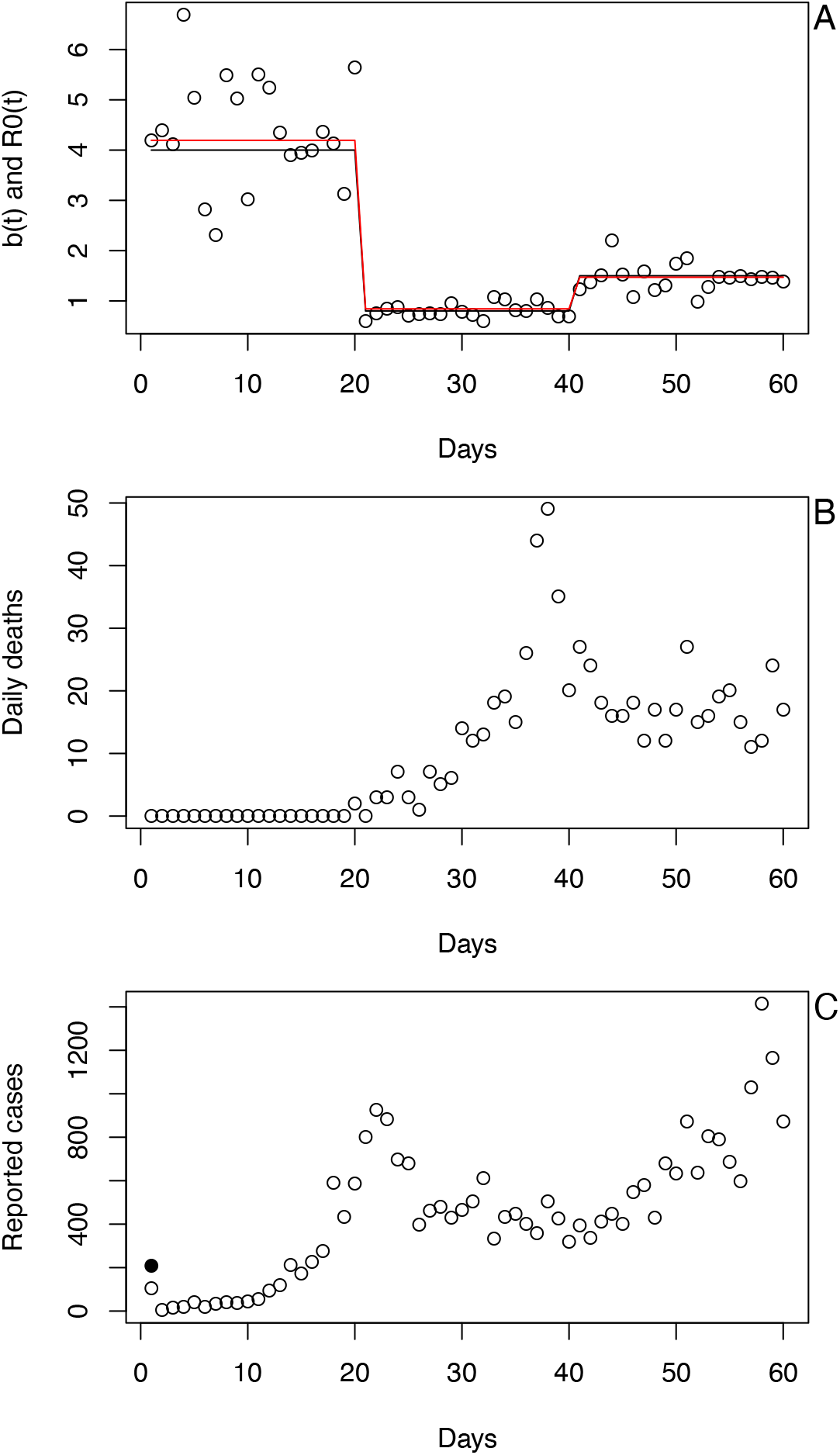
Example simulation from the process-based model. **(A)** Changes in the infection rate, *b*(*t*), are modeled as a step function (black line) with daily variation (points). R(*t*) (red line) tracks changes in *b*(*t*). **(B)** and **(C)** The number of deaths (B) and diagnosed cases (C) when the simulation is initiated with a single cohort of individuals, all infected on day 1 (solid black dot).

**Fig. S5.**
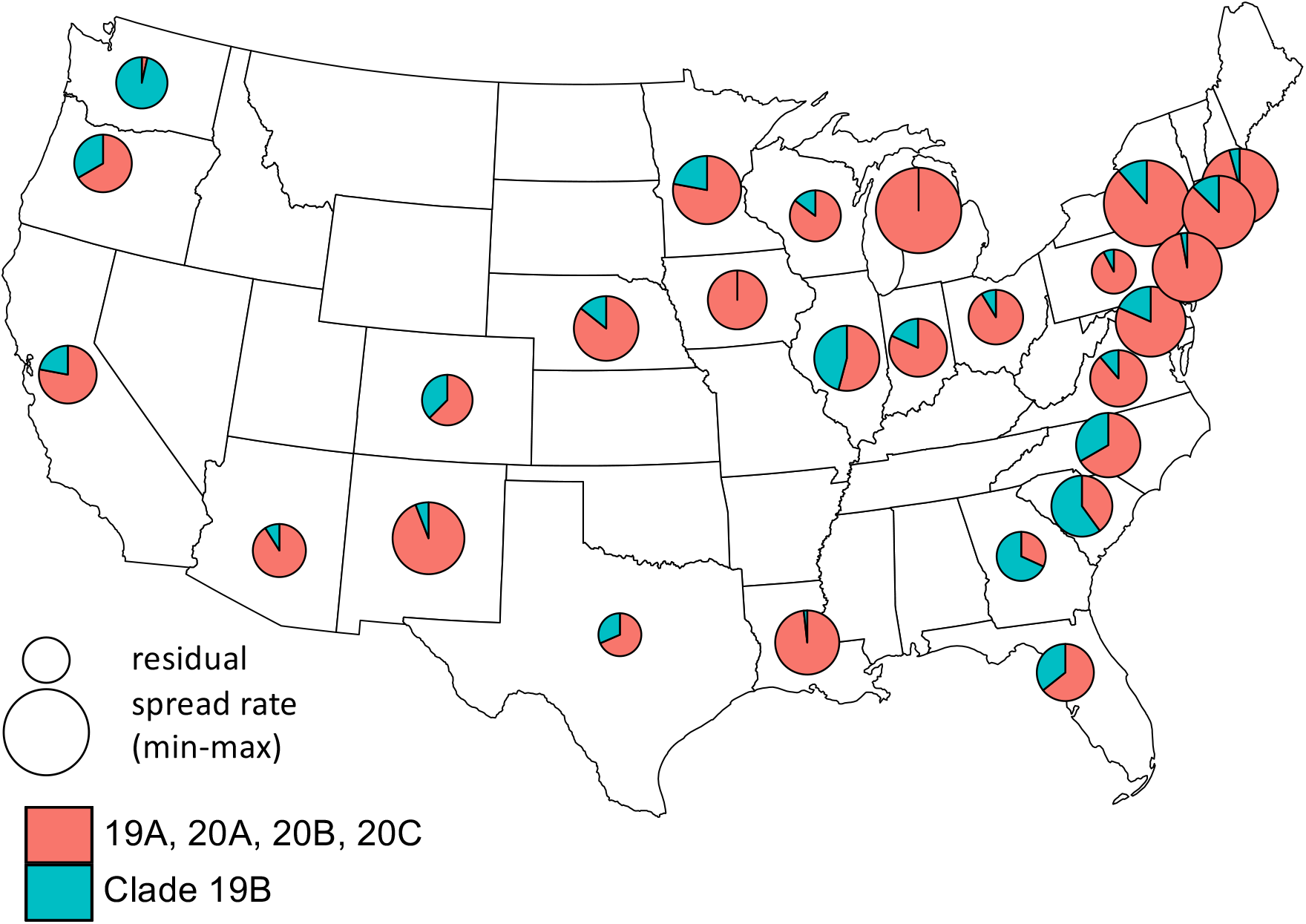
Spatial distribution of the 19B clade of SARS-CoV-2 at the outbreak onset among states. Pie charts give the proportion of samples in states collected within 30 days following the outbreak onset that are in the 19B clade (blue). The size of the pie is proportional to the residual values of *r*_0_ after removing the effects of the timing of outbreak onset, population size represented by the time series, and population density. For each state, we used the estimate of *r*_0_ corresponding to the county or county-aggregate that had the greatest number of deaths.

**Table S1.**
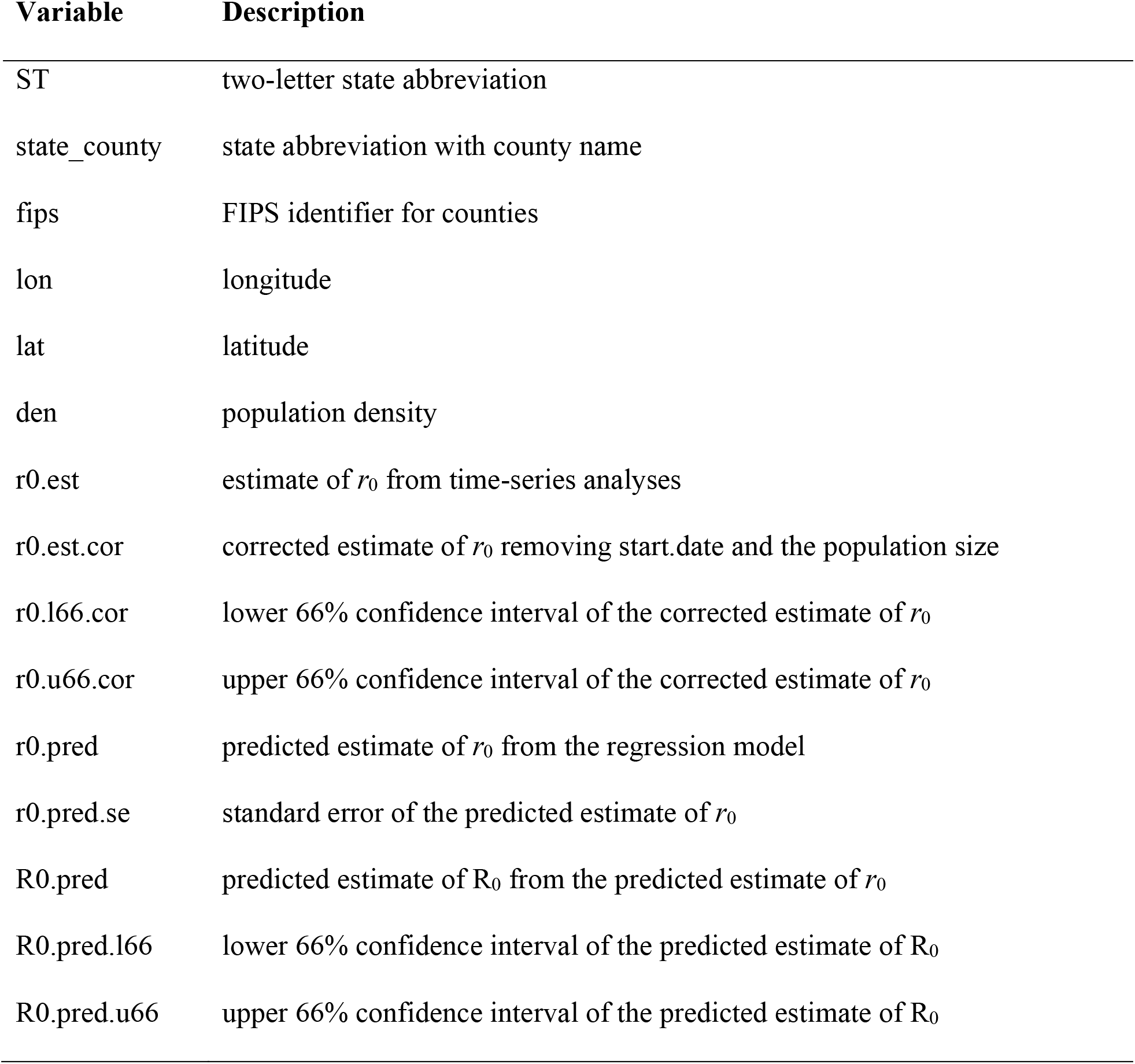
Separate spreadsheet giving the following variables for the 3109 counties in the conterminous USA.

| Variable | Description |
| --- | --- |
| ST | two-letter state abbreviation |
| state_county | state abbreviation with county name |
| fips | FIPS identifier for counties |
| lon | longitude |
| lat | latitude |
| den | population density |
| r0.est | estimate of $r_0$ from time-series analyses |
| r0.est.cor | corrected estimate of $r_0$ removing start.date and the population size |
| r0.l66.cor | lower 66% confidence interval of the corrected estimate of $r_0$ |
| r0.u66.cor | upper 66% confidence interval of the corrected estimate of $r_0$ |
| r0.pred | predicted estimate of $r_0$ from the regression model |
| r0.pred.se | standard error of the predicted estimate of $r_0$ |
| R0.pred | predicted estimate of $R_0$ from the predicted estimate of $r_0$ |
| R0.pred.l66 | lower 66% confidence interval of the predicted estimate of $R_0$ |
| R0.pred.u66 | upper 66% confidence interval of the predicted estimate of $R_0$ |

**Table S2.** Regression of the initial spread rate, *r*_0_, of COVID-19 against (i) the date of outbreak onset, (ii) total population size, (iii) population density, and (iv) the proportion of samples of SARS-CoV-2 containing the G614 mutation in the spike gene^10^. The estimates of *r*_0_ were for the county or county-aggregate with the greatest number of deaths in the state. All genetic samples were collected within 30 days following the onset of outbreak in a county. Twenty-eight states had five or more genetic samples, and only these states are included in the regression. Transforms of population size and density were selected to best-fit the data and satisfy linearity assumptions.

|  | **Coefficient** | **SE** | **t** | **P** |
| --- | --- | --- | --- | --- |
| **onset** | −0.0032 | 0.0013 | −2.42 | 0.024 |
| **log(size)** | 0.020 | 0.009 | 2.14 | 0.043 |
| **density<sup>1/4</sup>** | 0.012 | 0.005 | 2.28 | 0.033 |
| **G614** | 0.133 | 0.051 | 2.61 | 0.016 |

**Table S3.** Regression of the initial spread rate, *r*_0_, of COVID-19 against (i) the date of outbreak onset, (ii) total population size, (iii) population density, and (iv) the proportion of samples of SARS-CoV-2 in four of the five clades identified in the Nextstrain metadata^11^. The estimates of *r*_0_ were for the county or county-aggregate with the greatest number of deaths in the state. All genetic samples were collected within 30 days following the onset of outbreak in a county. Twenty-seven states had five or more genetic samples, and only these states are included in the regression. Transforms of population size and density were selected to best-fit the data and satisfy linearity assumptions.

|  | **Coefficient** | **SE** | **t** | **P** |
| --- | --- | --- | --- | --- |
| **onset** | -0.0032 | 0.0015 | -2.2 | 0.040 |
| **log(size)** | 0.019 | 0.011 | 1.82 | 0.085 |
| **density<sup>1/4</sup>** | 0.015 | 0.006 | 2.68 | 0.015 |
| **19A** | -0.050 | 0.095 | -0.53 | 0.60 |
| **19B** | -0.147 | 0.054 | -02.72 | 0.014 |
| **20A** | -0.031 | 0.057 | -0.54 | 0.59 |
| **20B** | 0.009 | 0.173 | 0.05 | 0.96 |

**Table S4.** Variables giving population characteristics^12-19^ that were included in the regression model to assess the importance of population density and spatial autocorrelation in the estimation of *r*_0_.

| **Variable** | **Description** |
| --- | --- |
| age | proportion of the population over 65 years old, 2011-2015 |
| adult obesity | incidence of adult obesity, 2015 |
| diabetes | incidence of adult diabetes, 2015 |
| education | percent bachelor's degree or higher, 2011-2015 |
| income | median earnings 2011-2015 |
| poverty | percentage people below federal poverty threshold, 2011-2015 |
| economic equality | Gini index, 2013-14 |
| race | percent White, non-Latino, 2015 |
| political leaning | proportion of votes cast for Donald Trump, 2016 |

**Table S5.** For 160 county and county-aggregates, regression of spread rate at the end of the time series, corresponding to 5 May, 2020, *r*(*t*_*end*_), against (i) the date of outbreak onset, (ii) total population size and (iii) population density, in which (iv) spatial autocorrelation is incorporated into the residual error. Transforms of population size and density were selected to best-fit the data and satisfy linearity assumptions. The coefficient column contains the estimate of the regression parameters with their associated t-tests; spatial autocorrelation is characterized by a range and nugget for regional and local sources of variation, and their joint significance is given by a likelihood ratio test. For the overall model, R^2^_pred_ = 0.40.

|  | **Coefficient** | **SE** | **t** | **P** | **partial $R^2_{pred}$** |
| --- | --- | --- | --- | --- | --- |
| **onset** | 0.0020 | 0.0003 | 6.18 | $< 10^{-8}$ | 0.15 |
| **log(size)** | 0.0093 | 0.0021 | 4.43 | $< 10^{-6}$ | 0.070 |
| **density<sup>1/4</sup>** | -0.0010 | 0.0014 | -0.68 | 0.50 | 0.003 |
| **space** | range = 0.24<br>nugget = 0 | | $\chi^2_2 = 13.89$ | 0.001 | 0.13 |

