## Supplementary material for "Estimating and explaining the spread of COVID-19 at the county level in the USA": Source code and compiled data: Ives&Bozzuto_Analysis_and_Figures_Rcode_20Oct20.R

```
library(usmap)
library(rr2)
library(nlme)
library(geosphere)

# Transmission (Li et al., 2020)
duration <- 25
age <- 1:duration

T0 <- 7.5
m <- T0
v <- 3.4^2

v/m^2
shape <- 2.35
gamma(1 + 2/shape)/gamma(1 + 1/shape)^2 - 1
scale <- m/gamma(1 + 1/shape)

pdf.trans <- dweibull(age, shape=shape, scale = scale)
pdf.trans <- pdf.trans/sum(pdf.trans)

##########################################################################################
#analyes with deaths forward and backwards
##########################################################################################

i.data <- "deaths"
d.for <- read.table(paste0("USA_counties_TRV.rev=FALSE_",i.data,"_sr.fixed.min=0.02_27May20.csv"), header=T, sep=",")
d.rev <- read.table(paste0("USA_counties_TRV.rev=TRUE_",i.data,"_sr.fixed.min=0.02_27May20.csv"), header=T, sep=",")

d.for <- d.for[!is.na(d.for$r0),]
d.rev <- d.rev[!is.na(d.rev$r0),]

par(mfrow=c(1,1), mai=c(.8,.8,.1,.1))
plot(d.for$r0.est ~ d.rev$r0.est)
lines(c(-1,1),c(-1,1))

par(mfrow=c(2,1))
hist(d.for$r0.Pr^.5)
hist(d.rev$r0.Pr^.5)

sr.df <- data.frame(state_county = d.for$state_county, d.for$sr, d.rev$sr)
sr.df[order(sr.df$state_county),]

##########################################################################################
# curate data
d.for[d.for$r0.est == min(d.for$r0.est, na.rm=T),]

d <- d.for
d[,10:42] <- (d.for[,10:42] + d.rev[,10:42])/2
d[d$state_county == "NY_Orange", 10:42] <- d.rev[d$state_county == "NY_Orange",10:42]

# add NY City fips (using fips for New York county * 10)
d$fips[d$state_county == "NY_New York City"] <- 360610

names(d)[names(d) == "abbr"] <- "ST"

##########################################################################################
d$state_county <- as.factor(d$state_county)
d$state_county <- droplevels(d$state_county)
d$start.date <- as.Date(d$start.date)

d <- d[order(d$state_county),]

dim(d)
d[d$count.max == min(d$count.max),]
levels(d$state_county)

d$R0 <- 1/(exp(-matrix(d$r0.est,ncol=1) %*% age) %*% pdf.trans)
d$l95 <- 1/(exp(-matrix(d$r0.l95,ncol=1) %*% age) %*% pdf.trans)
d$u95 <- 1/(exp(-matrix(d$r0.u95,ncol=1) %*% age) %*% pdf.trans)
d$l66 <- 1/(exp(-matrix(d$r0.l66,ncol=1) %*% age) %*% pdf.trans)
d$u66 <- 1/(exp(-matrix(d$r0.u66,ncol=1) %*% age) %*% pdf.trans)

##########################################################################################
# add county data
w1 <- read.csv("USA_county_data_13Oct20.csv", header=T, sep=",")
unique(w1$ST)

# missing county
# 46102 Oglala Lakota 
# 46113 SD_NA (Shannon)
w1[w1$fips == 46102,]

#remove unaggregated NYC boroughs (to be added back later)
NYC.fips <- c(36005, 36047, 36061, 36081, 36085)
dim(w1)
w1 <- w1[!is.element(w1$fips, NYC.fips),]
dim(w1)

d$fips[!is.element(d$fips, w1$fips)]

fips.list <- unique(d$fips)
fips.list <- fips.list[!is.na(fips.list)]

w1$thresh <- is.element(w1$fips, d$fips)

unique(d$ST)[!is.element(unique(d$ST), unique(w1$ST))]
unique(w1$ST)[!is.element(unique(w1$ST), unique(d$ST))]
# [1] "AK" "HI" "ID" "ME" "MT" "ND" "SD" "UT" "VT" "WV" "WY"

d[d$ST == "PR",]
d <- d[d$ST != "PR",]
d$ST <- as.factor(d$ST)

w1$ST <- as.factor(w1$ST)
nlevels(w1$ST)
nlevels(d$ST)

################
# where is NYC?
d[d$ST == "NY",1:4]

################
# w0 contains all the county info
w0 <- w1

# wu contains information for counties that with no estiamte of r0
wu <- w1
dim1 <- dim(wu)
wu <- wu[!is.element(wu$fips, d$fips),]
dim1 - dim(wu)
sum(wu$thresh)

################
# w contains information for counties with estimates of r0
w <- w1[1:nrow(d),]
w$state_county.w <- d$state_county
w$fips <- d$fips
w$ST <- d$ST
w$ST <- droplevels(w$ST)
nlevels(w$ST)

w$ST <- as.character(w$ST)
w0$ST <- as.character(w0$ST)
wu$ST <- as.character(wu$ST)

names(w)
for(i in 1:nrow(w)){
	if(!is.na(w$fips[i])) {
		w[i,-ncol(w)] <- w1[w1$fips == w$fips[i],]
	}else{
		ww <- w1[w1$ST == w$ST[i] & w1$thresh == F,]
		w[i, 1:4] <- NA
		w$ST[i] <- ww$ST[1]
		w[i, c(18, 37, 45)] <- colSums(ww[,c(18, 37, 45)], na.rm=T)
		w[i, c(5:17, 19:27, 32:36, 38:44, 47)] <- colSums(ww[,c(5:17, 19:27, 32:36, 38:44, 47)] * ww$Population.MA)/sum(ww$Population.MA)
	}
}

w$ST <- as.factor(w$ST)
w0$ST <- as.factor(w0$ST)

w[,c(1:3, 48:49)]

# merge info into d
d <- cbind(d, w)
dim(d)
d[,c("state_county", "state_county.w")]

d$pop <- d$Population.Census2019
d$den <- d$pop/d$area
d$den <- d$den/2.58999	

d[,c("state_county", "state_county.w", "den", "pop", "area")]

# include regions
unique(d$ST)
Southern.states <- c("AL","AR","FL","GA","KT","LA","MO","MS","NC","OK","SC","TN","TX","VA","WV")
Midwestern.states <- c("IA","IL","IN","KS","MI","MN","NE","OH","WI")
Western.states <- c("AZ","CA","CO","NM","OR","WA")
d$region <- "North"
d$region[is.element(d$ST, Southern.states)] <- "South" #red
d$region[is.element(d$ST, Midwestern.states)] <- "Midwest" #gray
d$region[is.element(d$ST, Western.states)] <- "West" #blue

write.table(d, "County data with r0.ests 13Oct20.csv", sep=',', row.names=F)

# make wu
wu$pop <- wu$Population.Census2019
wu$den <- wu$pop/wu$area
wu$den <- wu$den/2.58999	

col.list <- c("ST","fips", "state_county5","lat", "lon", "den")
ww <- wu[,col.list]
head(ww,20)

# missing county
# 46102 Oglala Lakota 
ww[ww$fips == 46102,]

write.table(ww, "County data without r0.ests 13Oct20.csv", sep=',', row.names=F)

##########################################################################################
# data descriptors
##########################################################################################

d <- read.csv("County data with r0.ests 13Oct20.csv")
d$state_county <- as.factor(d$state_county)
d$state <- as.factor(d$state)
d$ST <- as.factor(d$ST)
d$region <- as.factor(d$region)
d$start.date <- as.Date(d$start.date)

# number of aggregates
sum(grepl("_agg", d$state_county))

# number of states
nlevels(d$ST)

# median count.max
hist(exp(d$count.max))
median(exp(d$count.max))
exp(mean(d$count.max))

# proportion after 11 March
sum(d$start.date > as.Date("2020-03-11"))/nrow(d)

# last date
max(as.Date(d$end.date))

# first start.date
min(d$start.date)

# correlation between log(pop) and log(den)
cor(log(d$pop), log(d$den))

##########################################################################################
# Fig 1: plot r0
##########################################################################################

pdf(file="County Fig1 13Oct20.pdf", height=5, width=8)

	col.list <- c("gray","lightblue", "black", "blue", "black", "cyan")
	ds <- d[order(d$r0.est, decreasing = T),]
	ds$col <- 1
	ds$col[!ds$thresh] <- 2

	par(mfrow=c(1,1), mai=c(1,1,.1,.1))
	
	plot(1:nrow(ds), ds$r0.est, xaxt="n", xlab="Sorted counties", ylab = expression(paste("Estimated ", italic(r)[0])), ylim=c(-.08, .42), pch="", xlim=c(5, 155), cex.lab = 1.5)
	arrows(x0 = 1:nrow(ds), y0 = ds$r0.l95, x1 = 1:nrow(ds), y1 = ds$r0.u95, length = 0, col=col.list[ds$col + 2])
	arrows(x0 = 1:nrow(ds), y0 = ds$r0.l66, x1 = 1:nrow(ds), y1 = ds$r0.u66, length = 0, lwd=5, col=col.list[ds$col])
	points(1:nrow(ds), ds$r0.est, pch=15, col=col.list[ds$col + 4], cex=.7)
	lines(c(0,200), c(0,0), lty=2)
	
dev.off()

##########################################################################################
# version of R2_pred_gls
##########################################################################################
R2_pred.gls <- function(mod = NULL, mod.r = NULL) {
  y <- as.numeric(fitted(mod)+resid(mod))
  n <- mod$dims$N
#browser()  
  cormatrix <- nlme::corMatrix(mod$modelStruct$corStruct)
  if(!is.null(attr(mod$modelStruct$varStruct, 'weights'))){
    Vdiag <- 1/attr(mod$modelStruct$varStruct, 'weights')
    V <- diag(Vdiag) %*% cormatrix %*% diag(Vdiag)
  }else{
    V <- cormatrix
  }
  V <- sigma(mod)^2 * V

  R <- y - fitted(mod)
  
  Rhat <- matrix(0, nrow = n, ncol = 1)
  Rhat.var <- matrix(0, nrow = n, ncol = 1)
  for (j in 1:n) {
    r <- R[-j]
    VV <- V[-j, -j]
    iVV <- solve(VV)
    v <- as.matrix(V[j, -j], ncol=1)
    Rhat[j] <- t(v) %*% iVV %*% r
    Rhat.var[j] <- V[j,j] - t(v) %*% iVV %*% v
  }
  
  Yhat <- as.numeric(fitted(mod) + Rhat)
  SSE.pred <- var(y - Yhat)
  
  # reduced model
  if (class(mod.r) == "gls") {

  cormatrix <- nlme::corMatrix(mod.r$modelStruct$corStruct)
  if(!is.null(attr(mod.r$modelStruct$varStruct, 'weights'))){
    Vdiag <- 1/attr(mod.r$modelStruct$varStruct, 'weights')
    V.r <- diag(Vdiag) %*% cormatrix %*% diag(Vdiag)
  }else{
    V.r <- cormatrix
  }
  V.r <- sigma(mod.r)^2 * V.r
    R.r <- y - fitted(mod.r)
    Rhat.r <- matrix(0, nrow = n, ncol = 1)
    for (j in 1:n) {
      r.r <- R.r[-j]
      VV.r <- V.r[-j, -j]
      iVV.r <- solve(VV.r)
      v.r <- V.r[j, -j]
      Rhat.r[j] <- v.r %*% iVV.r %*% r.r
      
    }
    Yhat.r <- as.numeric(fitted(mod.r) + Rhat.r)
  }
  
  if (class(mod.r) == "lm") {
    Yhat.r <- stats::fitted(mod.r)
  }
  
  SSE.pred.r <- var(y - Yhat.r)
  
  return(list(R2 = 1 - SSE.pred/SSE.pred.r, Yhat = Yhat, Rhat = Rhat, Yhat.se = Rhat.var^.5))
}

##########################################################################################
# analyze den, pop, start.date
##########################################################################################
par(mfcol=c(2,2), mai=c(.8,.8,.4,.4))
plot(r0.est ~ log(den), data=d, xlab="Population density", ylab = "r0.est", cex.lab=1.1)	
plot(d$den^.5, d$r0.est, xlab="Population density", ylab = "r0.est", cex.lab=1.1)	
plot(d$den^.25, d$r0.est, xlab="Population density", ylab = "r0.est", cex.lab=1.1)	
plot(d$den^.1, d$r0.est, xlab="Population density", ylab = "r0.est", cex.lab=1.1)	

# take 1/4 power of density
d$denp25 <- d$den^.25

z <- gls(r0.est ~ denp25 + log(pop) + start.date, weights = varFixed(~r0.est.se^2), correlation = corExp(value=c(.1,.5),nugget = T, form=~lon + lat), data=d, method="ML")
z.date <- gls(r0.est ~ denp25 + log(pop), weights = varFixed(~r0.est.se^2), correlation = corExp(nugget = T, form=~lon + lat), data=d, method="ML")
z.size <- gls(r0.est ~ denp25 + start.date, weights = varFixed(~r0.est.se^2), correlation = corExp(nugget = T, form=~lon + lat), data=d, method="ML")
z.den <- gls(r0.est ~ log(pop) + start.date, weights = varFixed(~r0.est.se^2), correlation = corExp(nugget = T, form=~lon + lat), data=d, method="ML")
z.space <- lm(r0.est ~ denp25 + log(pop) + start.date, weights = 1/r0.est.se^2, data=d)
z.0 <- lm(r0.est ~ 1, weights = 1/r0.est.se^2, data=d)
summary(z)
# Generalized least squares fit by maximum likelihood
  # Model: r0.est ~ denp25 + log(pop) + start.date 
  # Data: d 
        # AIC       BIC   logLik
  # -669.7777 -648.2515 341.8889

# Correlation Structure: Exponential spatial correlation
 # Formula: ~lon + lat 
 # Parameter estimate(s):
    # range    nugget 
# 5.7085979 0.3315084 
# Variance function:
 # Structure: fixed weights
 # Formula: ~r0.est.se^2 

# Coefficients:
               # Value Std.Error   t-value p-value
# (Intercept) 34.70338  7.658159  4.531556       0
# denp25       0.00968  0.001681  5.762507       0
# log(pop)     0.02474  0.002773  8.923607       0
# start.date  -0.00191  0.000417 -4.588715       0

 # Correlation: 
           # (Intr) denp25 lg(pp)
# denp25     -0.027              
# log(pop)   -0.583  0.080       
# start.date -1.000  0.026  0.580

# Standardized residuals:
       # Min         Q1        Med         Q3        Max 
# -1.7015744 -0.3512854  0.4031302  1.0485977  3.3330219 

# Residual standard error: 1.188124 
# Degrees of freedom: 160 total; 156 residual
anova(z, z.space)
        # Model df       AIC       BIC   logLik   Test  L.Ratio p-value
# z           1  7 -669.7777 -648.2515 341.8889                        
# z.space     2  5 -600.9534 -585.5775 305.4767 1 vs 2 72.82436  <.0001


r2 <- c(R2_pred.gls(z, z.0)$R2, R2_lik(z, z.0))
r2 <- rbind(r2,c(R2_pred.gls(z, z.date)$R2, R2_lik(z, z.date)))
r2 <- rbind(r2,c(R2_pred.gls(z, z.size)$R2, R2_lik(z, z.size)))
r2 <- rbind(r2,c(R2_pred.gls(z, z.den)$R2, R2_lik(z, z.den)))
r2 <- rbind(r2,c(R2_pred.gls(z, z.space)$R2, R2_lik(z, z.space)))
print(r2, digits = 2)
# r2 0.70 0.62
   # 0.11 0.11
   # 0.36 0.31
   # 0.14 0.17
   # 0.48 0.37


##########################################################################################
# corrected r0 and R0
##########################################################################################

maxpop <- max(log(d$pop))
mindate <- as.Date("2020-03-11")

correction <- -coef(z)["log(pop)"]*(log(d$pop) - maxpop) - coef(z)["start.date"]*(d$start.date - mindate)
d$r0.est.cor <- d$r0.est + correction
d$r0.est.cor <- as.numeric(d$r0.est.cor)
plot(r0.est.cor ~ r0.est, data=d)
c(mean(d$r0.est), mean(d$r0.est.cor))
c(sd(d$r0.est), sd(d$r0.est.cor))

d$r0.l95.cor <- d$r0.l95 + correction
d$r0.u95.cor <- d$r0.u95 + correction
d$r0.l66.cor <- d$r0.l66 + correction
d$r0.u66.cor <- d$r0.u66 + correction

d$R0.cor <- 1/(exp(-matrix(d$r0.est.cor,ncol=1) %*% age) %*% pdf.trans)
d$l95.cor <- 1/(exp(-matrix(d$r0.l95.cor,ncol=1) %*% age) %*% pdf.trans)
d$u95.cor <- 1/(exp(-matrix(d$r0.u95.cor,ncol=1) %*% age) %*% pdf.trans)
d$l66.cor <- 1/(exp(-matrix(d$r0.l66.cor,ncol=1) %*% age) %*% pdf.trans)
d$u66.cor <- 1/(exp(-matrix(d$r0.u66.cor,ncol=1) %*% age) %*% pdf.trans)

d$death.max <- exp(d$count.max)
d$county <- substring(d$state_county, first=4)
d$county[d$county == "agg"] <- NA

##########################################################################################
# Fig 2 with corrected r0 
##########################################################################################
location <- d[,c("lon", "lat")]
names(location) <- c("log","lat")
dist <- distm(location)/1000
hist(dist)
max(dist)

n <- nrow(d)
i.r0.est <- matrix(d$r0.est.cor, n, n, byrow=F)
j.r0.est <- matrix(d$r0.est.cor, n, n, byrow=T)

breaks <- c(100*(0:10), 6000)

r0.dist <- data.frame(dist = breaks[-length(breaks)])
for(i in 1:(length(breaks)-1)){
	x1 <- i.r0.est[breaks[i] <= dist & dist < breaks[i+1] & !is.na(dist)] - mean(d$r0.est.cor)
	x2 <- j.r0.est[breaks[i] <= dist & dist < breaks[i+1] & !is.na(dist)] - mean(d$r0.est.cor)
	r0.dist$cor[i] <- mean(x1*x2)
	show(c(length(x1), r0.dist$cor[i]))
}
r0.dist$cor <- r0.dist$cor/sd(d$r0.est.cor)^2
r0.dist
plot(r0.dist)

################################
# remove distances > 1000
r0.dist <- r0.dist[-nrow(r0.dist),]
################################

d$region.num <- 1
d$region.num[d$region == "South"] <- 2
d$region.num[d$region == "West"] <- 4
d$region.num[d$region == "Midwest"] <- 5

    # range    nugget 
# 6.4145045 0.3488564 
range <- 6.4145045 
nugget <- 0.3488564  

# assuming 1 degree lat and lon = 100 km
range_km <- 100*range

r0.dist$p <- (1-nugget)*exp(-(r0.dist$dist/range_km))	

pdf("County Fig2 13Oct20.pdf", width=10, height=4.5)

	par(mfrow=c(1,2), mai=c(.9,.9,.4,.4), cex.lab = 1.4)
	
	plot(r0.est.cor ~ denp25, data=d, xlab=expression(paste("Population density (km"^"-2",")")), ylab = expression(paste("Corrected ", italic(r)[0])), ylim=c(0, .41), xaxt = "n", col=d$region.num, pch=d$region.num)	
	axis(1, labels = c(10, 100, 1000, 10000), at=c(10, 100, 1000, 10000)^.25)
	mtext("A", side=4, cex=1.8, las=1, padj=-7., adj=-.2)
	
	y <- r0.dist$cor
	dist.cat <- c("100", "200","300","400","500","600","700","800","900","1000")
	barplot(y, ylab=expression(paste("Correlation of corrected ", italic(r)[0])), xlab="Distance (km)", names.arg = dist.cat, ylim=c(-.05, .8))	
	lines(1.2*(1:10) - .6,r0.dist$p, col="green", lwd=2)
	box()
	mtext("B", side=4, cex=1.8, las=1, padj=-7., adj=-.2)
	
dev.off()

##########################################################################################
# Other variables
##########################################################################################

factor.list <- c("Elderly_Population.MA", "Obesity.MA", "Diabetes", "At_Least_Bachelor_Degree.MA", "Median_Earnings.AHDP", "Below_federal_poverty_threshold.MA", "Gini_Coefficient.MA", "White.AHDP", "pTrump")

for(i.factor in factor.list) {
	show(i.factor)
	d$dummy <- d[,i.factor]
	z.factor <- gls(r0.est ~ denp25 + log(pop) + start.date + dummy, weights = varFixed(~r0.est.se^2), correlation = corExp(nugget = T, form=~lon + lat), data=d, method="ML", na.action = na.omit)
	z.factor.space <- gls(r0.est ~ denp25 + log(pop) + start.date + dummy, weights = varFixed(~r0.est.se^2), data=d, method="ML", na.action = na.omit)
	
	show(summary(z.factor)$tTable)
	show(logLik(z.factor) - logLik(z.factor.space))
}

#########################################################################################
# Fig 4AB with corrected R0
##########################################################################################
mod <- gls(r0.est ~ log(pop) + denp25 + start.date, weights = varFixed(~r0.est.se^2), correlation = corExp(nugget = T, form=~lon + lat), data=d, method="ML")
mod.0 <- lm(r0.est ~ 1, weights = 1/r0.est.se^2, data=d)
summary(mod)

R2.mod <- R2_pred.gls(mod, mod.0)
d$Yhat <- R2.mod$Yhat
d$Yhat.se <- R2.mod$Yhat.se

maxpop <- max(log(d$pop))
mindate <- as.Date("2020-03-11")

correction <- -coef(z)["log(pop)"]*(log(d$pop) - maxpop) - coef(z)["start.date"]*(d$start.date - mindate)
correction <- as.numeric(correction)

d$r0.est.cor <- d$r0.est + correction
d$r0.pred <- d$Yhat + correction
d$r0.pred.se <- d$Yhat.se

d$r0.l95.cor <- d$r0.l95 + correction
d$r0.u95.cor <- d$r0.u95 + correction
d$r0.l66.cor <- d$r0.l66 + correction
d$r0.u66.cor <- d$r0.u66 + correction

d$R0.pred <- 1/(exp(-matrix(d$r0.pred,ncol=1) %*% age) %*% pdf.trans)
d$R0.cor <- 1/(exp(-matrix(d$r0.est.cor,ncol=1) %*% age) %*% pdf.trans)
d$l95.cor <- 1/(exp(-matrix(d$r0.l95.cor,ncol=1) %*% age) %*% pdf.trans)
d$u95.cor <- 1/(exp(-matrix(d$r0.u95.cor,ncol=1) %*% age) %*% pdf.trans)
d$l66.cor <- 1/(exp(-matrix(d$r0.l66.cor,ncol=1) %*% age) %*% pdf.trans)
d$u66.cor <- 1/(exp(-matrix(d$r0.u66.cor,ncol=1) %*% age) %*% pdf.trans)

summary(lm(r0.est.cor ~ r0.pred, data=d))
summary(lm(r0.est.cor ~ r0.pred, data=d))

# compare r0 estimates
colMeans(d[,c("r0.est","r0.est.cor", "r0.pred")])
d[,c("state_county", "r0.est","r0.est.cor", "r0.pred")]
d[,c("state_county", "lat","lon")]


##########################################################################################
# function pred.u for counties with no estimates of r0
##########################################################################################
pred.u <- function(mod, Vc, dat) {
	y <- as.numeric(fitted(mod)+resid(mod))
	n <- mod$dims$N

	cormatrix <- nlme::corMatrix(mod$modelStruct$corStruct)
	if(!is.null(attr(mod$modelStruct$varStruct, 'weights'))){
		Vdiag <- 1/attr(mod$modelStruct$varStruct, 'weights')
		V <- diag(Vdiag) %*% cormatrix %*% diag(Vdiag)
	}else{
		V <- cormatrix
	}
	V <- sigma(mod)^2 * V
	R <- y - fitted(mod)
	
	Vc <- sigma(mod)^2 * Vc

	n.m <- ncol(Vc)
	n.u <- nrow(Vc)
  
	Rhat <- matrix(NA, nrow = n.u, ncol = 1)
	Rhat.var <- matrix(NA, nrow = n.u, ncol = 1)

	iV <- solve(V)

	for(i in 1:n.u){
		v <- Vc[i,]
		Rhat[i] <- v %*% iV %*% R
		Rhat.var[i] <- sigma(mod)^2 - v %*% iV %*% v
	}

	Yhat <- coef(mod)["(Intercept)"] + coef(mod)["log(pop)"]*maxpop + coef(mod)["start.date"]*as.numeric(mindate) +  coef(mod)["denp25"]*dat$denp25 + Rhat

	Yhat.se <- Rhat.var^.5
	
    return(list(Yhat = Yhat, Yhat.se = Yhat.se, Rhat = Rhat))
}

#########################################################################################
# prediction of new values
#########################################################################################
wu <- read.csv("County data without r0.ests 13Oct20.csv")
wu$ST <- as.factor(wu$ST)
wu$denp25 <- wu$den^.25
summary(wu)

wu[wu$fips == 46103 & !is.na(wu$fips),]

state.aggs <- d$ST[is.na(d$county)]
length(state.aggs)
for(i.agg in state.aggs){
	pick1 <- wu$ST == i.agg
	pick2 <- d$ST == i.agg & is.na(d$county)
	wu$r0.est[pick1] <- d$r0.est[pick2]
	wu$r0.est.cor[pick1] <- d$r0.est.cor[pick2]
	wu$r0.l95.cor[pick1] <- d$r0.l95.cor[pick2]
	wu$r0.l66.cor[pick1] <- d$r0.l66.cor[pick2]
	wu$r0.u95.cor[pick1] <- d$r0.u95.cor[pick2]
	wu$r0.u66.cor[pick1] <- d$r0.u66.cor[pick2]
}
# check
summary(wu)
length(unique(wu$fips))
sum(is.element(wu$fips,d$fips))

n.m <- nrow(d)
n.u <- nrow(wu)

V <- matrix(NA, n.m, n.m)
for(i in 1:n.m) for(j in 1:n.m){
	dis <- sqrt((d$lat[i] - d$lat[j])^2 + (d$lon[i] - d$lon[j])^2)	
	V[i, j] <- (1-nugget)*exp(-dis/range)
}

Vc <- matrix(NA, n.u, n.m)
Dist <- matrix(NA, n.u, n.m)
for(i in 1:n.u) for(j in 1:n.m){
	dis <- sqrt((wu$lat[i] - d$lat[j])^2 + (wu$lon[i] - d$lon[j])^2)	
	Vc[i, j] <- (1-nugget)*exp(-dis/range)
	Dist[i, j] <- dis
}
min(Dist, na.rm=T)
max(Dist, na.rm=T)
mean(Dist, na.rm=T)
min(Vc, na.rm=T)
max(Vc, na.rm=T)
mean(Vc, na.rm=T)

# states with aggregates
range <- 6.4145045 
nugget <- 0.3488564  

for(i.agg in state.aggs){
	i <- which(ww$ST == i.agg)
	www <- wu[i,]
	dis <- 0
	for(ii in 1:(length(i)-1)) for(jj in (ii+1):length(i)){
		dis <- dis + sqrt((www$lat[ii] - www$lat[jj])^2 + (www$lon[ii] - www$lon[jj])^2)
	}
	dis <- dis * 2/(length(i)*(length(i)-1))
	show(dis)
	
	j <- which(d$ST == i.agg & is.na(d$county))
	Vc[i,j] <- (1-nugget)*exp(-dis/range)
}
# check
d[,1:6]
Vc[i,140:160]

mod.no.wt <- gls(r0.est ~ log(pop) + denp25 + start.date, correlation = corExp(nugget = T, form=~lon + lat), data=d, method="ML")
pred <- pred.u(mod = mod.no.wt, dat = wu, Vc=Vc) 
wu$r0.pred <- pred$Yhat
wu$r0.pred.se <- pred$Yhat.se

wu$R0.pred <- 1/(exp(-matrix(wu$r0.pred,ncol=1) %*% age) %*% pdf.trans)
wu$est <- F

d$est <- T
names(d)
names(wu)
col.list <- names(wu)
dw <- wu
is.element(names(wu),names(d))
dw <- rbind(dw, d[,col.list])
dw[is.na(dw$r0.est),]

dw[dw$fips == 46103 & !is.na(dw$fips),]

pdf("County Fig4AB 13Oct20.pdf", width=10, height=4.75)

	par(mfrow=c(1,2), mai=c(.9,.9,.4,.4), cex.lab = 1.4)
	
	plot(R0 ~ R0.pred, data=d, xlab = expression(paste("Predicted ", italic(R)[0])), ylab = expression(paste("Observed ", italic(R)[0])), xlim=c(.8, 8), ylim=c(.8, 8), log="xy", pch="")
	arrows(x0 = d$R0.pred, y0 = d$l66, x1 = d$R0.pred, y1 = d$u66, length = 0, lwd=2, col="gray")
	points(R0 ~ R0.pred, data=d)

	lines(c(.1,20), c(.1,20), col="red")

	plot(R0.cor ~ R0.pred, data=d, xlab = expression(paste("Predicted ", italic(R)[0])), ylab = expression(paste("Observed corrected ", italic(R)[0])), xlim=c(.8, 8), ylim=c(.8, 8), log="xy", pch="")
	arrows(x0 = d$R0.pred, y0 = d$l66.cor, x1 = d$R0.pred, y1 = d$u66.cor, length = 0, lwd=2, col="gray")
	points(R0.cor ~ R0.pred, data=d)

	lines(c(.1,20), c(.1,20), col="red")
	
dev.off()

dw$R0.est <- 1/(exp(-matrix(dw$r0.est,ncol=1) %*% age) %*% pdf.trans)
dw$R0.cor <- 1/(exp(-matrix(dw$r0.est.cor,ncol=1) %*% age) %*% pdf.trans)

# missing county
# 46102 Oglala Lakota 
# 46113 SD_NA (Shannon)
dw$fips[dw$fips > 46000 & dw$fips < 47000]

dw[dw$fips == 46102 & !is.na(dw$fips),]
dw[dw$fips == 46102 & !is.na(dw$fips),6:18] <- dw[dw$fips == 46103 & !is.na(dw$fips),6:18]
dw[dw$fips == 46102 & !is.na(dw$fips),]

# missing Weston county, WY
dw$fips[dw$fips > 56000 & dw$fips < 57000]
dw <- rbind(dw, dw[dw$fips == 56011 & !is.na(dw$fips),])
dw$fips[nrow(dw)] <- 56045

# missing NYC Boroughs
NYC.fips <- c(36005, 36047, 36061, 36081, 36085)
for(i in NYC.fips){
	dw <- rbind(dw, dw[dw$fips == 360610 & !is.na(dw$fips),])
	dw$fips[nrow(dw)] <- i
}
dw[is.element(dw$fips,NYC.fips),]
dw[dw$fips == 360610 & !is.na(dw$fips),]
dw <- dw[dw$fips != 360610 & !is.na(dw$fips),]

# Fig4C-E
library(usmap)
library(ggplot2)
require(gridExtra)

dw$logR0.est <- log(dw$R0.est)
dw$logR0.cor <- log(dw$R0.cor)
dw$logR0.pred <- log(dw$R0.pred)

map1 <- plot_usmap(regions = "counties", data = dw, values = "logR0.est", color = "white", include = .northeast_region) +   theme(legend.position = "right") +  scale_fill_continuous(low = "white", high = "red", label = scales::comma)
    
map2 <-  plot_usmap(regions = "counties", data = dw, values = "logR0.pred", color = "white", include = .northeast_region) +   theme(legend.position = "right") +  scale_fill_continuous(low = "white", high = "red", label = scales::comma)
gridExtra::grid.arrange(map1, map2, nrow=1)

plot_usmap(regions = "counties", data = dw, values = "logR0.pred", color = "white", exclude = c("AK","HI")) +   theme(legend.position = "right") +  scale_fill_continuous(low = "white", high = "red", label = scales::comma)

##########################################################################################
# Table S1 with corrected r0 and R0
##########################################################################################

dw$R0.pred <- 1/(exp(-matrix(dw$r0.pred,ncol=1) %*% age) %*% pdf.trans)
dw$R0.pred.l66 <- 1/(exp(-matrix(dw$r0.pred-dw$r0.pred.se,ncol=1) %*% age) %*% pdf.trans)
dw$R0.pred.u66 <- 1/(exp(-matrix(dw$r0.pred+dw$r0.pred.se,ncol=1) %*% age) %*% pdf.trans)

dw$R0.pressure <- 1 - 1/dw$R0.pred
dw$R0.pressure.l66 <- 1 - 1/dw$R0.pred.l66
dw$R0.pressure.u66 <- 1 - 1/dw$R0.pred.u66

aggregate(cbind(R0.pred, R0.pressure) ~ ST, data=dw, FUN=mean)
lapply(dw[,c("R0.pred", "R0.pressure")], FUN=max)
lapply(dw[,c("R0.pred", "R0.pressure")], FUN=min)

names(dw)
var.list <- c("ST", "state_county5", "fips", "lon", "lat", "den", "r0.est", "r0.est.cor", "r0.l66.cor", "r0.u66.cor", "r0.pred", "r0.pred.se", "R0.pred", "R0.pred.l66",  "R0.pred.u66")
is.element(var.list,names(dw))

dw <- dw[order(dw$fips),]
dw <- dw[dw$ST != "HI" & dw$ST !="AK",]

write.table(dw[,var.list], file="Ives&Bozzuto Table S1 13Oct20.csv", sep=",", row.names=F)


##########################################################################################
# analysis of r1
##########################################################################################
z.1 <- gls(r1.est ~ log(den) + log(pop) + start.date, weights = varFixed(~r1.est.se^2), correlation = corExp(nugget = T, form=~lon + lat), data=d, method="ML")
z.1.date <- gls(r1.est ~ log(den) + log(pop), weights = varFixed(~r1.est.se^2), correlation = corExp(nugget = T, form=~lon + lat), data=d, method="ML")
z.1.size <- gls(r1.est ~ log(den) + start.date, weights = varFixed(~r1.est.se^2), correlation = corExp(nugget = T, form=~lon + lat), data=d, method="ML", control=glsControl(opt="optim"))
z.1.den <- gls(r1.est ~ log(pop) + start.date, weights = varFixed(~r1.est.se^2), correlation = corExp(nugget = T, form=~lon + lat), data=d, method="ML")
z.1.space <- lm(r1.est ~ log(den) + log(pop) + start.date, weights = 1/r1.est.se^2, data=d)
z.1.0 <- lm(r1.est ~ 1, weights = 1/r1.est.se^2, data=d)

summary(z.1)
# Generalized least squares fit by maximum likelihood
  # Model: r1.est ~ log(den) + log(pop) + start.date 
  # Data: d 
        # AIC       BIC   logLik
  # -742.3945 -720.8683 378.1973

# Correlation Structure: Exponential spatial correlation
 # Formula: ~lon + lat 
 # Parameter estimate(s):
       # range       nugget 
# 2.421732e-01 1.850456e-08 
# Variance function:
 # Structure: fixed weights
 # Formula: ~r1.est.se^2 

# Coefficients:
                # Value Std.Error   t-value p-value
# (Intercept) -36.98990  5.984076 -6.181388  0.0000
# log(den)     -0.00098  0.001433 -0.683234  0.4955
# log(pop)      0.00934  0.002108  4.430005  0.0000
# start.date    0.00201  0.000326  6.175763  0.0000

 # Correlation: 
           # (Intr) lg(dn) lg(pp)
# log(den)   -0.001              
# log(pop)   -0.552  0.181       
# start.date -1.000 -0.001  0.548

# Standardized residuals:
       # Min         Q1        Med         Q3        Max 
# -2.6682847 -0.8347536 -0.1598857  0.5753825  2.5812095 

# Residual standard error: 0.7389096 
# Degrees of freedom: 160 total; 156 residual
anova(z.1, z.1.space)
          # Model df       AIC       BIC   logLik   Test  L.Ratio p-value
# z.1           1  7 -742.3945 -720.8683 378.1973                        
# z.1.space     2  5 -732.5007 -717.1248 371.2503 1 vs 2 13.89385   0.001

# fit with pglmm_compare to use R2_pred
R2_pred.gls(z.1, z.1.0)$R2
R2_pred.gls(z.1, z.1.date)$R2
R2_pred.gls(z.1, z.1.size)$R2
R2_pred.gls(z.1, z.1.den)$R2
R2_pred.gls(z.1, z.1.space)$R2
# [1] 0.399044
# [1] 0.1513265
# [1] 0.06986333
# [1] 0.003091567
# [1] 0.1266602

##########################################################################################
##########################################################################################
# Analysis of GISAID genetic clades
##########################################################################################
##########################################################################################
library(nlme)


#########################################
# Using county-level data
d <- read.csv("County data with r0.ests 13Oct20.csv", header=T, sep=",")
d$start.date <- as.Date(d$start.date)

d$ST <- as.factor(d$ST)
d$count.max <- exp(d$count.max)

summary(d)
dim(d)

d$select <- FALSE
for(i.ST in levels(d$ST)){
	pick <- (d$ST == i.ST) & !is.na(d$fips)
	if(any(pick)){
		d$select[which(pick)[d$count.max[pick] == max(d$count.max[pick])]] <- TRUE
	}else{
		d$select[d$ST == i.ST] <- TRUE
	}
}
d[,c(1:5, ncol(d))]
sum(d$select)

d <- d[d$select,]

#########################################
# Input strains from GISAID metadata
#########################################

w <- read.csv("GISAID metadata.csv", header=T)
names(w)
w <- w[w$country_exposure == "USA",]
dim(w)
#[1] 11488    26

w$Collection.Date <- as.Date(w$date, format="%m/%d/%Y")
min(w$Collection.Date, na.rm=T)
max(w$Collection.Date, na.rm=T)

w$division_exposure[w$division_exposure == "Washington DC"] <- "District of Columbia"

table(w$GISAID_clade)
table(w$division_exposure)

unique(d$state[!is.element(d$state, w$division_exposure)])
unique(w$division_exposure[!is.element(w$division_exposure, d$state)])

state.list <- unique(d$state[is.element(d$state, w$division_exposure)])
length(state.list)
sum(is.element(unique(w$division_exposure), state.list))
sum(is.element(unique(d$state), state.list))

state.list <- sort(state.list)
state.list
x <- data.frame(state = state.list)
for(i.state in x$state){
	ww <- w[w$division_exposure == i.state,]
	start.date <- d$start.date[i.state == d$state]
	ww <- ww[ww$Collection.Date <= 30 + start.date,]
	ww <- ww[!is.na(ww$GISAID_clade),]
	
	x$G[x$state == i.state] <- sum(ww$GISAID_clade == "G")
	x$GH[x$state == i.state] <- sum(ww$GISAID_clade == "GH")
	x$GR[x$state == i.state] <- sum(ww$GISAID_clade == "GR")
	x$L[x$state == i.state] <- sum(ww$GISAID_clade == "L")
	x$S[x$state == i.state] <- sum(ww$GISAID_clade == "S")
	x$O[x$state == i.state] <- sum(ww$GISAID_clade == "O")
	x$V[x$state == i.state] <- sum(ww$GISAID_clade == "V")
}

# remove states with few genotypes
x <- x[rowSums(x[,2:8]) >= 5,]
x
dim(x)

#########################################
# Analysis of genetic data
#########################################

x[,2:8] <- x[,2:8]/rowSums(x[,2:8])

# PCA
summary(pca <- princomp(x[,2:7], cor = FALSE, scores = TRUE))
biplot(pca)
colnames(pca$scores) <- paste0("c",colnames(pca$scores))
x <- cbind(x, pca$scores)

x[,9:14] <- x[,9:14]/rowSums(x[,9:14])
summary(pca.g <- princomp(x[,9:14], cor = FALSE, scores = TRUE))
biplot(pca.g)
colnames(pca.g$scores) <- paste0("g",colnames(pca.g$scores))
x <- cbind(x, pca.g$scores)
x

rowSums(x[,2:8])

dim(x)
dim(d)
d <- d[is.element(d$state, x$state),]
dim(d)

d <- cbind(d, x[match(d$state, x$state),])
d[,c(1:4, 73)]
d$start.date <- d$start.date - min(d$start.date)
d$den25 <- d$den^.25

# Spike position 614 (D614G): G, GR, GH

colMeans(x[,2:8])
summary(lm(r0.est ~ start.date + log(pop) + den25 + G + GH + GR + L + S + O + V, data=d, weights=1/d$r0.est.se^2))

d$G.clade <- d$G + d$GH + d$GR
summary(lm(r0.est ~ start.date + log(pop) + den25 + G.clade, data=d, weights=1/d$r0.est.se^2))
# Weighted Residuals:
    # Min      1Q  Median      3Q     Max 
# -2.3998 -0.6299 -0.1585  0.7380  2.6015 

# Coefficients:
             # Estimate Std. Error t value Pr(>|t|)  
# (Intercept) -0.268677   0.156470  -1.717   0.0994 .
# start.date  -0.003170   0.001316  -2.409   0.0244 *
# log(pop)     0.020095   0.009377   2.143   0.0429 *
# den25        0.011753   0.005165   2.275   0.0325 *
# G.clade      0.133202   0.051019   2.611   0.0156 *
# ---
# Signif. codes:  0 ‘***’ 0.001 ‘**’ 0.01 ‘*’ 0.05 ‘.’ 0.1 ‘ ’ 1

# Residual standard error: 1.335 on 23 degrees of freedom
# Multiple R-squared:  0.5925,	Adjusted R-squared:  0.5217 
# F-statistic: 8.362 on 4 and 23 DF,  p-value: 0.0002563


summary(gls(r0.est ~ log(pop) + den25 + start.date + G.clade, weights = varFixed(~r0.est.se^2), correlation = corExp(nugget = T, form=~lon + lat), data=d, method="ML"))
summary(gls(r0.est ~ log(pop) + den25 + start.date, weights = varFixed(~r0.est.se^2), correlation = corExp(nugget = T, form=~lon + lat), data=d, method="ML"))

# significance of spatial autocorrelation
z.gls <- gls(r0.est ~ log(pop) + den25 + start.date + G.clade, weights = varFixed(~r0.est.se^2), correlation = corExp(nugget = T, form=~lon + lat), data=d, method="ML")
z.lm <- lm(r0.est ~ start.date + log(pop) + den25 + G.clade, data=d, weights=1/d$r0.est.se^2)
anova(z.gls, z.lm)
      # Model df       AIC       BIC   logLik   Test      L.Ratio p-value
# z.gls     1  8 -86.17466 -75.51702 51.08733                            
# z.lm      2  6 -90.17466 -82.18143 51.08733 1 vs 2 5.500027e-09       1

# significance of spatial autocorrelation without strains
z.gls <- gls(r0.est ~ log(pop) + den25 + start.date, weights = varFixed(~r0.est.se^2), correlation = corExp(nugget = T, form=~lon + lat), data=d, method="ML")
z.lm <- lm(r0.est ~ start.date + log(pop) + den25, data=d, weights=1/d$r0.est.se^2)
anova(z.gls, z.lm)
      # Model df       AIC       BIC   logLik   Test   L.Ratio p-value
# z.gls     1  7 -81.09607 -71.77064 47.54804                         
# z.lm      2  5 -84.90692 -78.24590 47.45346 1 vs 2 0.1891554  0.9098

summary(lm(r0.est ~ start.date + log(pop) + den25 + cComp.1 + cComp.2 + cComp.3, data=d, weights=1/d$r0.est.se^2))
# lm(formula = r0.est ~ start.date + log(pop) + den25 + cComp.1 + 
    # cComp.2 + cComp.3, data = d, weights = 1/d$r0.est.se^2)

# Weighted Residuals:
    # Min      1Q  Median      3Q     Max 
# -2.2990 -0.6548 -0.1810  0.6766  2.5040 

# Coefficients:
             # Estimate Std. Error t value Pr(>|t|)  
# (Intercept) -0.163882   0.160899  -1.019   0.3200  
# start.date  -0.002983   0.001408  -2.119   0.0462 *
# log(pop)     0.018908   0.009630   1.963   0.0630 .
# den25        0.012353   0.005307   2.328   0.0300 *
# cComp.1      0.102814   0.041430   2.482   0.0216 *
# cComp.2     -0.024265   0.066582  -0.364   0.7192  
# cComp.3      0.008152   0.107066   0.076   0.9400  
# ---
# Signif. codes:  0 ‘***’ 0.001 ‘**’ 0.01 ‘*’ 0.05 ‘.’ 0.1 ‘ ’ 1

# Residual standard error: 1.375 on 21 degrees of freedom
# Multiple R-squared:  0.6051,	Adjusted R-squared:  0.4923 
# F-statistic: 5.363 on 6 and 21 DF,  p-value: 0.001708


# Map
library(usmap)
library(ggplot2)
library(scatterpie)

# residuals
mod <- lm(r0.est ~ start.date + log(pop) + den25, data=d, weights=1/d$r0.est.se^2)
d$resid <- mod$resid

# use average of county ln/lat
dat <- read.csv("USA_county_data_13Oct20.csv", header=T, sep=",")
for(i.state in unique(d$ST)){
	d$state.lon[d$ST == i.state] <- mean(dat$lon[dat$ST == i.state], na.rm=T)
	d$state.lat[d$ST == i.state] <- mean(dat$lat[dat$ST == i.state], na.rm=T)
}
             
# using GISAID
dmap <- d[,c("state.lon","state.lat","resid","G.clade")]
names(dmap)[1:2] <- c("lon","lat")
dmap <- usmap_transform(dmap)

dmap$scale <- .5 + .5*(dmap$resid - min(dmap$resid))/(max(dmap$resid) - min(dmap$resid))
dmap$scale <- 1.5*10^5*dmap$scale
dmap$col2 <- 1 - dmap$G.clade
dmap$col1 <- dmap$G.clade

plot_usmap(exclude = c("AK","HI")) + geom_scatterpie(data = dmap, aes(x = lon.1, y = lat.1, r = scale), cols = c("col1","col2"))

pdf("County Fig3 19Oct20.pdf", width=12, height=9)             
plot_usmap(exclude = c("AK","HI")) + geom_scatterpie(data = dmap, aes(x = lon.1, y = lat.1, r = scale), cols = c("col1","col2"), sorted_by_radius = T) + guides(fill = guide_legend(title = "", title.position = "top")) + theme(legend.position = "none")
dev.off()    

##########################################################################################
##########################################################################################
# Analysis of NextStrain genetic clades
##########################################################################################
##########################################################################################
library(nlme)

#########################################
# Using county-level data
d <- read.csv("County data with r0.ests 13Oct20.csv", header=T, sep=",")
d$start.date <- as.Date(d$start.date)

d$ST <- as.factor(d$ST)
d$count.max <- exp(d$count.max)

summary(d)
dim(d)

d$select <- FALSE
for(i.ST in levels(d$ST)){
	pick <- (d$ST == i.ST) & !is.na(d$fips)
	if(any(pick)){
		d$select[which(pick)[d$count.max[pick] == max(d$count.max[pick])]] <- TRUE
	}else{
		d$select[d$ST == i.ST] <- TRUE
	}
}
d[,c(1:5, ncol(d))]
sum(d$select)

d <- d[d$select,]

#########################################
# Input strains from nextstrain

w <- read.csv("nextstrain_ncov_north-america_metadata.csv", header=T)
names(w)
w <- w[w$Country.of.exposure == "USA",]
dim(w)
w$Collection.Date <- as.Date(w$Collection.Date)

unique(w$Clade)
table(w$Clade)

unique(w$GISAID.Clade)
table(w$GISAID.Clade)

unique(d$state[!is.element(d$state, w$Admin.Division)])
unique(w$Admin.Division[!is.element(w$Admin.Division, d$state)])

state.list <- unique(d$state[is.element(d$state, w$Admin.Division)])
length(state.list)
sum(is.element(unique(w$Admin.Division), state.list))
sum(is.element(unique(d$state), state.list))

state.list <- sort(state.list)

x <- data.frame(state = state.list)
for(i.state in x$state){
	ww <- w[w$Admin.Division == i.state,]
	start.date <- d$start.date[i.state == d$state]
	ww <- ww[ww$Collection.Date <= 30 + start.date,]
	x$c19A[x$state == i.state] <- sum(ww$Clade == "19A")
	x$c19B[x$state == i.state] <- sum(ww$Clade == "19B")
	x$c20A[x$state == i.state] <- sum(ww$Clade == "20A")
	x$c20B[x$state == i.state] <- sum(ww$Clade == "20B")
	x$c20C[x$state == i.state] <- sum(ww$Clade == "20C")
	
	x$G[x$state == i.state] <- sum(ww$GISAID.Clade == "G")
	x$GH[x$state == i.state] <- sum(ww$GISAID.Clade == "GH")
	x$GR[x$state == i.state] <- sum(ww$GISAID.Clade == "GR")
	x$L[x$state == i.state] <- sum(ww$GISAID.Clade == "L")
	x$S[x$state == i.state] <- sum(ww$GISAID.Clade == "S")
	x$O[x$state == i.state] <- sum(ww$GISAID.Clade == "O")
	x$V[x$state == i.state] <- sum(ww$GISAID.Clade == "V")
}

# for comparison
w <- w[is.element(w$Admin.Division, state.list),]
x19A <- aggregate(Clade ~ Admin.Division, data=w, FUN=function(x) sum(x == "19A"))
x19B <- aggregate(Clade ~ Admin.Division, data=w, FUN=function(x) sum(x == "19B"))
x20A <- aggregate(Clade ~ Admin.Division, data=w, FUN=function(x) sum(x == "20A"))
x20B <- aggregate(Clade ~ Admin.Division, data=w, FUN=function(x) sum(x == "20B"))
x20C <- aggregate(Clade ~ Admin.Division, data=w, FUN=function(x) sum(x == "20C"))
xx <- cbind(x19A, x19B[,2], x20A[,2], x20B[,2], x20C[,2])
names(xx)[2:6] <- c("c19A", "c19B", "c20A", "c20B", "c20C")
cbind(xx, x)

# remove states with few genotypes
x <- x[rowSums(x[,2:6]) >= 5,]

# PCA
x[,2:6] <- x[,2:6]/rowSums(x[,2:6])
summary(pca <- princomp(x[,2:5], cor = FALSE, scores = TRUE))
biplot(pca)
colnames(pca$scores) <- paste0("c",colnames(pca$scores))
x <- cbind(x, pca$scores)

x[,7:13] <- x[,7:13]/rowSums(x[,7:13])
summary(pca.g <- princomp(x[,7:12], cor = FALSE, scores = TRUE))
biplot(pca.g)
colnames(pca.g$scores) <- paste0("g",colnames(pca.g$scores))
x <- cbind(x, pca.g$scores)
x

dim(x)
dim(d)
d <- d[is.element(d$state, x$state),]
dim(d)

d <- cbind(d, x[match(d$state, x$state),])
d[,c(1:4, 73)]
d$start.date <- d$start.date - min(d$start.date)
d$den25 <- d$den^.25

summary(lm(r0.est ~ start.date + log(pop) + den25 + c19A + c19B + c20A + c20B, data=d, weights=1/d$r0.est.se^2))
# lm(formula = r0.est ~ start.date + log(pop) + den25 + c19A + 
    # c19B + c20A + c20B, data = d, weights = 1/d$r0.est.se^2)

# Weighted Residuals:
    # Min      1Q  Median      3Q     Max 
# -2.5569 -0.5169 -0.0667  0.8741  2.2495 

# Coefficients:
             # Estimate Std. Error t value Pr(>|t|)  
# (Intercept) -0.134389   0.170521  -0.788   0.4404  
# start.date  -0.003198   0.001453  -2.201   0.0403 *
# log(pop)     0.019436   0.010687   1.819   0.0848 .
# den25        0.015163   0.005657   2.681   0.0148 *
# c19A        -0.050379   0.095321  -0.529   0.6033  
# c19B        -0.146810   0.054017  -2.718   0.0137 *
# c20A        -0.031054   0.057364  -0.541   0.5946  
# c20B         0.008826   0.172505   0.051   0.9597  
# ---
# Signif. codes:  0 ‘***’ 0.001 ‘**’ 0.01 ‘*’ 0.05 ‘.’ 0.1 ‘ ’ 1

# Residual standard error: 1.381 on 19 degrees of freedom
# Multiple R-squared:  0.6395,	Adjusted R-squared:  0.5068 
# F-statistic: 4.816 on 7 and 19 DF,  p-value: 0.002935
        
```
