## Supplementary material for "Estimating and explaining the spread of COVID-19 at the county level in the USA": Source code and compiled data: Ives&Bozzuto_Testing_TVR_EpiEstim_13Oct20.R

```
#setwd("D:/beastdata/arives/Covid-19 County/")

library(mgcv)
library(parallel)
library(EpiEstim)
source("tvr_27May20.R")


dir.name <- "test_TVR_EpiEstim_2Sep20"
dir.create(dir.name)
pdf_path <- paste0("./", dir.name,"/")


###########################################################
# find parameters for simulations
delta <- 0.5
nrep <- 20
i.count <- 0

df <- data.frame(i.data = rep("deaths", nrep*2), r.shape = NA, death.max = NA, Tstart = NA, duration = NA, b0 = NA)
#for(i.data in c("deaths","cases")) for(r.shape in c("step.step","geometric")){
for(i.data in "deaths") for(r.shape in c("step.step","geometric")){
	
	if(i.data == "deaths"){
		analysis.threshold <- 50
		initiate.threshold <- 1
		length.threshold <- 20
		range0.threshold <- 1
		k <- 10
		moving.ave <- 1
		I.init <- 200
	}else{
		analysis.threshold <- 300
		initiate.threshold <- 5
		length.threshold <- 20
		range0.threshold <- 10
		k <- 10
		moving.ave <- 1
		I.init <- 200
	}
	
	
	i.rep <- 0
	for(i in 1:nrep){
		
		i.rep <- i.rep + 1
		i.count <- i.count + 1
		
		if(r.shape == "step.step"){
			b0.max <- 8
			b0.min <- 1.5
			
			b0.max <- log(b0.max)
			b0.min <- log(b0.min)
					
			b0 = b0.min + (i.rep - 1)*(b0.max - b0.min)/(nrep - 1)
			b0 <- exp(b0)
			b0.max <- exp(b0.max)
			b0.min <- exp(b0.min)
			
			Tmax.min <- 55
			Tmax.max <- 150			
			ppower <- 4
			Tmax <- round(Tmax.min + (Tmax.max - Tmax.min)*((b0 - b0.max)/(b0.min - b0.max))^ppower)

			b1 = .8
			b2 = 1.5
			b <- c(rep(b0, ceiling(Tmax/3)), rep(b1, ceiling(Tmax/3)), rep(b2, ceiling(Tmax/3)))
		}
		
		if(r.shape == "geometric"){
			b0.max <- 8
			b0.min <- 1.5
			
			b0.max <- log(b0.max)
			b0.min <- log(b0.min)
					
			b0 = b0.min + (i.rep - 1)*(b0.max - b0.min)/(nrep - 1)
			b0 <- exp(b0)
			b0.max <- exp(b0.max)
			b0.min <- exp(b0.min)
			
			Tmax.min <- 55
			Tmax.max <- 150
			ppower <- 4
			Tmax <- round(Tmax.min + (Tmax.max - Tmax.min)*((b0 - b0.max)/(b0.min - b0.max))^ppower)
			
			b1.max <- .5
			b1.min <- .8
			b1 = b1.min + (i.rep - 1)*(b1.max - b1.min)/(nrep - 1)
			
			b <- b0 * (b1/b0)^((1:Tmax - 1)/(Tmax - 1))			
		}
		
		covid.sim <- covid19_sim(b = b, initx = 1, I.init = I.init, surv = .98, se = .2, sm.incub = .2, sm.mort = .2, show.fig = F)
		
		#cases = row 1, deaths = row 2
		if(i.data == "deaths"){
			x.sim <- covid.sim$X[2,]
			day.lag <- 18
		}else{
			x.sim <- covid.sim$X[1,-1]
			day.lag <- 5
		}
			
		Tstart <- min(which(x.sim >= initiate.threshold))
		X <- x.sim[Tstart:length(x.sim)]

		x <- X
		# x <- x + delta		
		x[x==0] <- delta
		x <- log(x)
			
		day <- 1:length(x)
		dat <- data.frame(x = x, day = day)		

		mod.gamm <- try(gamm(x ~ s(day, k = k), data = dat, correlation = corARMA(form = ~ day, p=1, q=1), method = "ML", weights = varExp(value = -1, form = ~fitted(.))), silent = TRUE)
		if(inherits(mod.gamm, "try-error")){
			next
		}
		x.fitted.gamm = as.numeric(predict(mod.gamm$gam, newdata=dat))
		if(!any(x.fitted.gamm >= log(range0.threshold))) next
		range0 <- sort(which(x.fitted.gamm >= log(range0.threshold)))[1]

		range1 <- length(x)
		range <- range0:range1
		
		Tstart <- Tstart + range0
		
		df$i.data[i.count] <- i.data
		df$r.shape[i.count] <- r.shape
		df$death.max[i.count] <- max(exp(x), na.rm=T)
		df$Tstart[i.count] <-Tstart
		df$duration[i.count] <- length(x)
		df$b0[i.count] <- b0
	}
}		
par(mfrow=c(2,2))
for(r.shape in c("step.step","geometric")) {
	plot(death.max ~ b0, data=df[df$r.shape == r.shape,])
	plot(duration ~ b0, data=df[df$r.shape == r.shape,])
}

###########################################################
# set up pdf.trans for EpiEstim
duration <- 25
age <- 1:duration

T0 <- 7.5
m <- T0
v <- 3.4^2

v/m^2
shape <- 2.35
gamma(1 + 2/shape)/gamma(1 + 1/shape)^2 - 1
scale <- m/gamma(1 + 1/shape)

pdf.trans <- dweibull(age, shape=shape, scale = scale)
pdf.trans <- pdf.trans/sum(pdf.trans)


pdf.trans[1] <- 0
pdf.trans <- pdf.trans/sum(pdf.trans)

###########################################################
# set up parameters for TVR
annealing <- T
se.fixed <- 0
sr.fixed <- NA
sr.fixed.min <- 0.02
sr0.fixed <- 0
sm.start <- .5
sr.start <- .01

delta <- .5

moving.ave <- 1
i.0 <- (moving.ave+1)/2


nboot <- 100
nrep <- 30

###########################################################
# simulations
trial <- 2
	
start.flag <- T

#for(i.data in c("deaths","cases")) for(r.shape in c("step.step","geometric")){
for(i.data in "deaths") for(r.shape in c("step.step","geometric")){
	
	if(i.data == "deaths"){
		analysis.threshold <- 100
		initiate.threshold <- 2
		length.threshold <- 8
		range0.threshold <- 1
		k <- 10
		moving.ave <- 1
		I.init <- 200
	}else{
		analysis.threshold <- 3000
		initiate.threshold <- 20
		length.threshold <- 10
		range0.threshold <- 30
		k <- 10
		moving.ave <- 1
		I.init <- 200
	}
	
	i.rep <- 0
	for(i in 1:nrep){
		
		i.rep <- i.rep + 1
		
		if(r.shape == "step.step"){
			b0.max <- 8
			b0.min <- 1.5
			
			b0.max <- log(b0.max)
			b0.min <- log(b0.min)
					
			b0 = b0.min + (i.rep - 1)*(b0.max - b0.min)/(nrep - 1)
			b0 <- exp(b0)
			b0.max <- exp(b0.max)
			b0.min <- exp(b0.min)
			
			Tmax.min <- 55
			Tmax.max <- 150			
			ppower <- 4
			Tmax <- round(Tmax.min + (Tmax.max - Tmax.min)*((b0 - b0.max)/(b0.min - b0.max))^ppower)

			b1 = .8
			b2 = 1.5
			b <- c(rep(b0, ceiling(Tmax/3)), rep(b1, ceiling(Tmax/3)), rep(b2, ceiling(Tmax/3)))
		}
		
		if(r.shape == "geometric"){
			b0.max <- 8
			b0.min <- 1.5
			
			b0.max <- log(b0.max)
			b0.min <- log(b0.min)
					
			b0 = b0.min + (i.rep - 1)*(b0.max - b0.min)/(nrep - 1)
			b0 <- exp(b0)
			b0.max <- exp(b0.max)
			b0.min <- exp(b0.min)
			
			Tmax.min <- 55
			Tmax.max <- 150
			ppower <- 4
			Tmax <- round(Tmax.min + (Tmax.max - Tmax.min)*((b0 - b0.max)/(b0.min - b0.max))^ppower)
			
			b1.max <- .5
			b1.min <- .8
			b1 = b1.min + (i.rep - 1)*(b1.max - b1.min)/(nrep - 1)
			
			b <- b0 * (b1/b0)^((1:Tmax - 1)/(Tmax - 1))			
		}
		
		show(b)

		covid.sim <- covid19_sim(b = b, initx = 1, I.init = I.init, surv = .98, se = .2, sm.incub = .2, sm.mort = .2, show.fig = F)
		
		#cases = row 1, deaths = row 2
		if(i.data == "deaths"){
			x.sim <- covid.sim$X[2,]
			day.lag <- 18
		}else{
			x.sim <- covid.sim$X[1,-1]
			day.lag <- 5
		}
			
		Tstart <- min(which(x.sim >= initiate.threshold))
		X <- x.sim[Tstart:length(x.sim)]

		x <- X
		# x <- x + delta		
		x[x==0] <- delta
		x <- log(x)
			
		day <- 1:length(x)
		dat <- data.frame(x = x, day = day)		

		mod.gamm <- try(gamm(x ~ s(day, k = k), data = dat, correlation = corARMA(form = ~ day, p=1, q=1), method = "ML", weights = varExp(value = -1, form = ~fitted(.))), silent = TRUE)
		if(inherits(mod.gamm, "try-error")){
			next
		}
		x.fitted.gamm = as.numeric(predict(mod.gamm$gam, newdata=dat))
		if(!any(x.fitted.gamm >= log(range0.threshold))) next
		range0 <- sort(which(x.fitted.gamm >= log(range0.threshold)))[1]

		range1 <- length(x)
		range <- range0:range1
		
		Tstart <- Tstart + range0
		Tstart <- max(Tstart, day.lag + 1)
		
		if(length(range) <= length.threshold) next
		
		x <- x[range]
		dat <- dat[range,]
		X <- X[range]
		
			
		##############################################
		# EpiEstim

		dat.Epi <- data.frame(I = X)
		z <- try(estimate_R(dat.Epi, 
				method="non_parametric_si",
				config = make_config(list(si_distr = pdf.trans))), silent=TRUE)
		if(!inherits(z, "try-error")){

			df <- data.frame(i.data = NA, r.shape = NA, method = NA, sr.fixed = sr.fixed, sr = NA, death.max = NA, duration = NA, Tstart = NA, b.Tstart = NA, r.Tstart = NA, r.Tend = NA, r0 = NA, r0.est = NA, r0.u95 = NA, r0.u66 = NA, r0.l66 = NA, r0.l95 = NA, R0 = NA, R0.u95 = NA, R0.l95 = NA)
			
			plot(z)
			
			df$r.shape <- r.shape
			df$i.data <- i.data
	
			df$death.max <- max(x.sim)
			df$Tstart <- Tstart
			df$duration <- length(x)
			
			df$b.Tstart <- covid.sim$b[Tstart-day.lag+1]
			df$r.Tstart <- covid.sim$r[Tstart-day.lag+1]
			df$r.Tend <- covid.sim$r[Tstart-day.lag+length(x)]
			
			df$method <- "EpiEstim"
			
			df$R0 <- z$R$'Mean(R)'[1]
			df$R0.l95 <- z$R$'Quantile.0.025(R)'[1]
			df$R0.u95 <- z$R$'Quantile.0.975(R)'[1]
			
			if(start.flag == T) {
				write.table(df, paste0("test_TVR_EpiEstim_trial=", trial,"_28May20.csv"), sep=',', row.names = F)
				start.flag <- F
			}else{
				write.table(df, paste0("test_TVR_EpiEstim_trial=", trial,"_28May20.csv"), sep=',', row.names = F, append = T, col.names = F)
			}
		}
		# changing interval
		# n <- length(X)
		# t_start <- 2:(n-2)		
		# t_end <- 4:n		
		# z <- try(estimate_R(dat, 
				# method="non_parametric_si",
				# config = make_config(list(si_distr = pdf.trans, t_start=t_start, t_end=t_end))), silent=TRUE)
		

	
		##############################################
		# tvr
		mod.for <- try(tvr(x, se.fixed = se.fixed, sr.fixed = sr.fixed, sr0.fixed = sr0.fixed,
				sm.start = sm.start, sr.start = sr.start,  
				annealing = annealing),, silent = T)
		if(!inherits(mod.for, "try-error") & mod.for$sr < sr.fixed.min){				
				mod.for <- try(tvr(x, se.fixed = 0, sr.fixed = sr.fixed.min, sr0.fixed = sr0.fixed,
				sm.start = sm.start, sr.start = sr.start,  
				annealing = annealing), silent = T)
		}
		
		if(!inherits(mod.for, "try-error")){

			df <- data.frame(i.data = NA, r.shape = NA, method = NA, sr.fixed = sr.fixed, sr = NA, death.max = NA, duration = NA, Tstart = NA, b.Tstart = NA, r.Tstart = NA, r.Tend = NA, r0 = NA, r0.est = NA, r0.u95 = NA, r0.u66 = NA, r0.l66 = NA, r0.l95 = NA, R0 = NA, R0.u95 = NA, R0.l95 = NA)
			
			show(mod.for)
			
			###### set sr to its fitted value for bootstrapping
			mod.for$sr.fixed <- mod.for$sr
			mod.for$sm.start <- mod.for$sm
			b.for <- boot_tvr(mod.for, nboot=nboot, annealing = annealing, moving.ave = moving.ave, r.order=c(1,0,0))
			
			df$r.shape <- r.shape
			df$i.data <- i.data
			
			df$death.max <- max(x.sim)
			df$Tstart <- Tstart
			df$duration <- length(x)
			
			df$b.Tstart <- covid.sim$b[Tstart-day.lag+1]
			df$r.Tstart <- covid.sim$r[Tstart-day.lag+1]
			df$r.Tend <- covid.sim$r[Tstart-day.lag+length(x)]
			
			df$method <- "TVR.for"
			
			df$sr <- mod.for$sr
	
			df$r0 <- mod.for$r.fitted[i.0]
			df$r0.est <- mean(b.for$r[,i.0], na.rm=T)
			df$r0.u95 <- b.for$r.upper95[i.0]
			df$r0.u66 <- b.for$r.upper66[i.0]
			df$r0.l66 <- b.for$r.lower66[i.0]
			df$r0.l95 <- b.for$r.lower95[i.0]
		
			df$R0 <- 1/(exp(-matrix(df$r0.est,ncol=1) %*% age) %*% pdf.trans)
			df$R0.u95 <- 1/(exp(-matrix(df$r0.u95,ncol=1) %*% age) %*% pdf.trans)
			df$R0.l95 <- 1/(exp(-matrix(df$r0.l95,ncol=1) %*% age) %*% pdf.trans)

			if(start.flag == T) {
				write.table(df, paste0("test_TVR_EpiEstim_trial=", trial,"_28May20.csv"), sep=',', row.names = F)
				start.flag <- F
			}else{
				write.table(df, paste0("test_TVR_EpiEstim_trial=", trial,"_28May20.csv"), sep=',', row.names = F, append = T, col.names = F)
			}
		}
		
		##############################################
		# tvr rev = T
		mod.rev <- try(tvr(x, rev = T, se.fixed = se.fixed, sr.fixed = sr.fixed, sr0.fixed = sr0.fixed,
				sm.start = sm.start, sr.start = sr.start,  
				annealing = annealing), silent = T)
		
		if(!inherits(mod.rev, "try-error") & mod.rev$sr < sr.fixed.min){				
				mod.rev <- try(tvr(x, rev = T, se.fixed = 0, sr.fixed = sr.fixed.min, sr0.fixed = sr0.fixed,
				sm.start = sm.start, sr.start = sr.start,  
				annealing = annealing), silent = T)
		}
		if(!inherits(mod.rev, "try-error")){

			df <- data.frame(i.data = NA, r.shape = NA, method = NA, sr.fixed = sr.fixed, sr = NA, death.max = NA, duration = NA, Tstart = NA, b.Tstart = NA, r.Tstart = NA, r.Tend = NA, r0 = NA, r0.est = NA, r0.u95 = NA, r0.u66 = NA, r0.l66 = NA, r0.l95 = NA, R0 = NA, R0.u95 = NA, R0.l95 = NA)
			
			show(mod.rev)
			
			###### set sr to its fitted value for bootstrapping
			mod.rev$sr.fixed <- mod.rev$sr
			mod.rev$sm.start <- mod.rev$sm
			b.rev <- boot_tvr(mod.rev, nboot=nboot, annealing = annealing, moving.ave = moving.ave, r.order=c(1,0,0))
			
			df$r.shape <- r.shape
			df$i.data <- i.data
			
			df$death.max <- max(x.sim)
			df$Tstart <- Tstart
			df$duration <- length(x)
			
			df$b.Tstart <- covid.sim$b[Tstart-day.lag+1]
			df$r.Tstart <- covid.sim$r[Tstart-day.lag+1]
			df$r.Tend <- covid.sim$r[Tstart-day.lag+length(x)]

			df$method <- "TVR.rev"
			
			df$sr <- mod.for$sr
	
			df$r0 <- mod.rev$r.fitted[i.0]
			df$r0.est <- mean(b.rev$r[,i.0], na.rm=T)
			df$r0.u95 <- b.rev$r.upper95[i.0]
			df$r0.u66 <- b.rev$r.upper66[i.0]
			df$r0.l66 <- b.rev$r.lower66[i.0]
			df$r0.l95 <- b.rev$r.lower95[i.0]
		
			df$R0 <- 1/(exp(-matrix(df$r0.est,ncol=1) %*% age) %*% pdf.trans)
			df$R0.u95 <- 1/(exp(-matrix(df$r0.u95,ncol=1) %*% age) %*% pdf.trans)
			df$R0.l95 <- 1/(exp(-matrix(df$r0.l95,ncol=1) %*% age) %*% pdf.trans)

			if(start.flag == T) {
				write.table(df, paste0("test_TVR_EpiEstim_trial=", trial,"_28May20.csv"), sep=',', row.names = F)
				start.flag <- F
			}else{
				write.table(df, paste0("test_TVR_EpiEstim_trial=", trial,"_28May20.csv"), sep=',', row.names = F, append = T, col.names = F)
			}
		}
		
		
		pdf(file = paste0(pdf_path, "test_TVR_shape=",r.shape,"_",i.data,"_sr.fixed.min=",sr.fixed.min, "_rep=",i.rep,"_28May20.pdf"), width = 9, height = 8)
		
			par(mfcol=c(2,2), mai = c(.1,1,1,.1))
			
			mod <- mod.for
			b <- b.for
			
			plot(exp(x), xlab="", ylab = "Number", xaxt = "n")
			lines(exp(mod$X.fitted))
			
			par(mai = c(1,1,.1,.1))
			plot(mod$r.fitted, type = "l", lty=1, col="black", ylab = "r", xlab = "", ylim=c(-.2,.4), lwd = 2)
			
			TT <- length(x) - (moving.ave-1)/2
			polygon(c(i.0:TT, TT:i.0), c(b$r.upper95[i.0:TT], rev(b$r.lower95[i.0:TT])), col="lightgray", border=NA)
			polygon(c(i.0:TT, TT:i.0), c(b$r.upper66[i.0:TT], rev(b$r.lower66[i.0:TT])), col="darkgray", border=NA)
			lines(mod$r.fitted)
			lines(c(0,length(x)), c(0,0), lty=2)
			
			lines(covid.sim$r[-(1:(Tstart-day.lag))], col="red")
		
			par(mai = c(.1,1,1,.1))

			mod <- mod.rev
			b <- b.rev
			
			plot(exp(x), xlab="", ylab = "Number", xaxt = "n")
			lines(exp(mod$X.fitted))
			
			par(mai = c(1,1,.1,.1))
			plot(mod$r.fitted, type = "l", lty=1, col="black", ylab = "r", xlab = "", ylim=c(-.2,.4), lwd = 2)
			
			TT <- length(x) - (moving.ave-1)/2
			polygon(c(i.0:TT, TT:i.0), c(b$r.upper95[i.0:TT], rev(b$r.lower95[i.0:TT])), col="lightgray", border=NA)
			polygon(c(i.0:TT, TT:i.0), c(b$r.upper66[i.0:TT], rev(b$r.lower66[i.0:TT])), col="darkgray", border=NA)
			lines(mod$r.fitted)
			lines(c(0,length(x)), c(0,0), lty=2)
			
			lines(covid.sim$r[-(1:(Tstart-day.lag))], col="red")
		
		dev.off()
		
	}
}

###############################################################################
# analyze output
###############################################################################
# set up pdf.trans for EpiEstim
duration <- 25
age <- 1:duration

T0 <- 7.5
m <- T0
v <- 3.4^2

v/m^2
shape <- 2.35
gamma(1 + 2/shape)/gamma(1 + 1/shape)^2 - 1
scale <- m/gamma(1 + 1/shape)

pdf.trans <- dweibull(age, shape=shape, scale = scale)
pdf.trans <- pdf.trans/sum(pdf.trans)


pdf.trans[1] <- 0
pdf.trans <- pdf.trans/sum(pdf.trans)

###########################################################
# set up parameters for TVR
annealing <- T
se.fixed <- 0
sr.fixed <- NA
sr.fixed.min <- 0.02
sr0.fixed <- 0
sm.start <- .5
sr.start <- .01

delta <- .5

moving.ave <- 1
i.0 <- (moving.ave+1)/2

nboot <- 100
nrep <- 30

###########################################################
df <- read.table("test_TVR_EpiEstim_trial=2_28May20.csv", sep=',', header = T)

########################################################
# all types
par(mfrow=c(4,2), mai=c(.8,.8,.4,.1))

for(i.method in c("EpiEstim", "TVR.for", "TVR.rev")) for(r.shape in c("step.step","geometric")){	
	
	d <- df[df$method == i.method & df$r.shape == r.shape,]
	
	plot(R0 ~ b.Tstart, data=d, xlab="True R0", ylab = "Est R0", ylim=c(.4,20), main=i.method, log="xy")
	arrows(x0 = d$b.Tstart, y0 = d$R0.l95, x1 = d$b.Tstart, y1 = d$R0.u95, length = 0, lwd=1, col="black")
	points(R0 ~ b.Tstart, data=d, pch=19)
	lines(c(.1,10), c(.1,10), lty=2)
}
for(r.shape in c("step.step","geometric")) {	
	
	i.method <- "TVR.for"
	d <- df[df$method == i.method & df$r.shape == r.shape,]
	i.method <- "TVR.rev"
	d.rev <- df[df$method == i.method & df$r.shape == r.shape,]
	d$r0.est <- (d$r0.est + d.rev$r0.est)/2
	d$r0.u95 <- (d$r0.u95 + d.rev$r0.u95)/2
	d$r0.l95 <- (d$r0.l95 + d.rev$r0.l95)/2
	
	d$R0 <- 1/(exp(-matrix(d$r0.est,ncol=1) %*% age) %*% pdf.trans)
	d$R0.u95 <- 1/(exp(-matrix(d$r0.u95,ncol=1) %*% age) %*% pdf.trans)
	d$R0.l95 <- 1/(exp(-matrix(d$r0.l95,ncol=1) %*% age) %*% pdf.trans)
	
	plot(R0 ~ b.Tstart, data=d, xlab="True R0", ylab = "Est R0", ylim=c(.4,20), main="TVR", log="xy")
	arrows(x0 = d$b.Tstart, y0 = d$R0.l95, x1 = d$b.Tstart, y1 = d$R0.u95, length = 0, lwd=1, col="black")
	points(R0 ~ b.Tstart, data=d, pch=19)
	lines(c(.1,10), c(.1,10), lty=2)
}
```
