## Supplementary material for "Estimating and explaining the spread of COVID-19 at the county level in the USA": Source code and compiled data: Ives&Bozzuto_Time_series_analysis_13Oct20.R

```
library(doParallel)
registerDoParallel(cores=4)

library(mgcv)
library(parallel)
source("tvr_27May20.R")


###########################################################
# compute r FUNCTION
###########################################################

compute_r <- function(x, d, annealing, se.fixed, sr.fixed, sr.fixed.min, sr0.fixed, sm.start, sr.start, delta, k, moving.ave, nboot){

	i.data <- x$data
	i.rev <- x$rev
		
	dir.name <- paste0("USA_counties_TVR.rev=",i.rev,"_", i.data,"_28May20")
	dir.create(dir.name)
	pdf_path <- paste0("./", dir.name,"/")

	if(i.data == "deaths"){
		range0.threshold <- 1
		initiate.threshold <- 1

	}else{
		range0.threshold <- 10
		initiate.threshold <- 5
	}
	###########################################################
	
	state_counties <- unique(d$state_county)
	
	df <- data.frame(state_county = state_counties, fips = NA, state = as.factor(1), abbr = as.factor(1), start.date = as.Date("2020-01-22"), end.date = as.Date("2020-01-22"), duration = NA, count.max = NA, sr = NA, r0 = NA, r0.Pr = NA, r0.ave3 = NA, r0.ave5 = NA, r0.est = NA, r0.est.se = NA, r0.est.ave3 = NA, r0.est.ave5 = NA, r0.u95 = NA, r0.u66 = NA, r0.l66 = NA, r0.l95 = NA, r1 = NA, r1.Pr = NA, r1.est = NA, r1.est.se = NA, r1.u95 = NA, r1.u66 = NA, r1.l66 = NA, r1.l95 = NA, rmax = NA, rmax.est = NA, rmax.u95 = NA, rmax.u66 = NA, rmax.l66 = NA, rmax.l95 = NA, rmin = NA, rmin.est = NA, rmin.u95 = NA, rmin.u66 = NA, rmin.l66 = NA, rmin.l95 = NA)
	levels(df$state) <- levels(d$state)
	levels(df$abbr) <- levels(d$abbr)
	
	fits <- data.frame(state_county = as.factor(1), type = as.factor(1), date = as.Date(d$date[1]), val = rep(NA, as.numeric(max(d$date) - min(d$date))+1))
	
	levels(fits$state_county) <- levels(d$state_county)
	levels(fits$type) <- c("x", "x.fitted", "r.fitted", "Pr.fitted", "r.se", "r.u95", "r.u66", "r.l66", "r.l95")
	
	start.flag <- T
	i.rep <- 0
	for(i.county in state_counties){
		i.rep <- i.rep + 1
		
		if(d$thresh[d$state_county == i.county][1]){
			
			# show(i.county)
	
			if(i.data == "deaths"){
				dd <- d[d$state_county == i.county, c("deaths", "date")]
				names(dd) <- c("y","date")
			}else{
				dd <- d[d$state_county == i.county, c("cases", "date")]
				names(dd) <- c("y","date")
			}	
			
			dd <- dd[order(dd$date),]
			
			dat.dif <- c(0, dd$y[-1] - dd$y[-length(dd$y)])
			
			# remove negative changes in cummulative cases
			dd$y[dat.dif < 0] <- NA
			dat.dif <- c(0, dd$y[-1] - dd$y[-length(dd$y)])
			
			Tstart <- min(which(dat.dif >= initiate.threshold))
			x <- dat.dif[Tstart:length(dat.dif)]
			x[x <= 0] <- delta
			x <- log(x)
			
			day <- 1:length(x)
			dat <- data.frame(x = x, day = day)		
			dat$date <- dd$date[-(1:(Tstart-1))]
			
			while(is.na(x[1])) {
				x <- x[-1]
				dat <- dat[-1,]
			}
			while(is.na(x[length(x)])) {
				x <- x[-length(x)]
				dat <- dat[-nrow(dat),]
			}
				
			# assuming a delay of 8 days for cases and 18 days for death
			if(i.data == "deaths"){
				dat$date <- dat$date - 18
			}else{
				dat$date <- dat$date - 8
			}	
	
			mod.gamm <- try(gamm(x ~ s(day, k = k), data = dat, correlation = corARMA(form = ~ day, p=1, q=1), method = "ML", weights = varExp(value = -1, form = ~fitted(.))), silent = TRUE)
			if(inherits(mod.gamm, "try-error")){
				show(i.county)
				next
			}
			x.fitted.gamm = as.numeric(predict(mod.gamm$gam, newdata=dat))
#			if(!any(x.fitted.gamm >= log(range0.threshold))) next
			range0 <- sort(which(x.fitted.gamm >= log(range0.threshold)))[1]
	
			range1 <- length(x)
			range <- range0:range1
			
			x <- x[range]
			dat <- dat[range,]
	
			while(is.na(x[1])) {
				x <- x[-1]
				dat <- dat[-1,]
			}
			while(is.na(x[length(x)])) {
				x <- x[-length(x)]
				dat <- dat[-nrow(dat),]
			}
	
			mod <- tvr(x, rev = i.rev, se.fixed = 0, sr.fixed = sr.fixed, sr0.fixed = sr0.fixed,
					sm.start = sm.start, sr.start = sr.start,  
					annealing = annealing)
			if(mod$sr < sr.fixed.min){				
					mod <- tvr(x, rev = i.rev, se.fixed = 0, sr.fixed = sr.fixed.min, sr0.fixed = sr0.fixed,
					sm.start = sm.start, sr.start = sr.start,  
					annealing = annealing)
			}
			
			###### set sr to its fitted value for bootstrapping
			mod$sr.fixed <- mod$sr
			mod$sm.start <- mod$sm
			b <- boot_tvr(mod, nboot=nboot, annealing = annealing, moving.ave = moving.ave)
	
			x.fitted = mod$X.fitted
			r.fitted <- mod$r.fitted
			
			if(moving.ave > 1){
				rr <- r.fitted
				if(any(is.na(rr))) rr[is.na(rr)] <- approx(rr, xout = which(is.na(rr)))$y
				r.fitted <- filter(rr, rep(1/moving.ave,moving.ave))
			}
						
			i.0 <- (moving.ave+1)/2
	
			png(file = paste0(pdf_path, "USA_county_TRV.rev=",i.rev,"_",i.data,"_",i.county,"_sr.fixed.min=",sr.fixed.min,"_27May20.png"), width = 480, height = 800)
				par(mfrow=c(2,1), mai = c(.1,1,1,.1))
				
				plot(exp(x), xlab="", ylab = "Number", xaxt = "n", col=1+as.numeric(weekdays(dat$date)=="Sunday"))
				lines(exp(x.fitted))
				
				par(mai = c(1,1,.1,.1))
				plot(r.fitted, type = "l", lty=1, col="black", ylab = "r(t)", xlab = "", ylim=c(-.2,.4), lwd = 2, xaxt = "n")
				
				TT <- length(x) - (moving.ave-1)/2
				polygon(c(i.0:TT, TT:i.0), c(b$r.upper95[i.0:TT], rev(b$r.lower95[i.0:TT])), col="lightgray", border=NA)
				polygon(c(i.0:TT, TT:i.0), c(b$r.upper66[i.0:TT], rev(b$r.lower66[i.0:TT])), col="darkgray", border=NA)
				lines(r.fitted)
				lines(c(0,length(x)), c(0,0), lty=2)
			
				lmax <- length(r.fitted)
				at.ticks <- round(1 + (0:5)/5 * (lmax - 1))
				axis(side = 1, at = at.ticks, labels = format(dat$date[at.ticks], "%d-%b"), las = 2)
			dev.off()
			
			df$state_county[i.rep] <- i.county
			df$fips[i.rep] <- d$fips[d$state_county == i.county][1]
			df$state[i.rep] <- d$state[d$state_county == i.county][1]
			df$abbr[i.rep] <- d$abbr[d$state_county == i.county][1]
			
			df$sr[i.rep] <- mod$sr
			df$count.max[i.rep] <- max(dat$x, na.rm=T)
			df$start.date[i.rep] <- dat$date[1]
			df$end.date[i.rep] <- dat$date[length(x)]
			df$duration[i.rep] <- length(x)
			
			df$r0[i.rep] <- r.fitted[i.0]
			df$r0.Pr[i.rep] <- mod$Pr[i.0]
			df$r0.ave3[i.rep] <- mean(r.fitted[i.0:(i.0+2)], na.rm=T)
			df$r0.ave5[i.rep] <- mean(r.fitted[i.0:(i.0+4)], na.rm=T)
					
			df$r0.est[i.rep] <- mean(b$r[,i.0], na.rm=T)
			df$r0.est.se[i.rep] <- b$r.se[i.0]
			df$r0.est.ave3[i.rep] <- mean(b$r[,i.0:(i.0+2)], na.rm=T)
			df$r0.est.ave5[i.rep] <- mean(b$r[,i.0:(i.0+4)], na.rm=T)
					
			df$r0.u95[i.rep] <- b$r.upper95[i.0]
			df$r0.u66[i.rep] <- b$r.upper66[i.0]
			df$r0.l66[i.rep] <- b$r.lower66[i.0]
			df$r0.l95[i.rep] <- b$r.lower95[i.0]
			
			df$r1[i.rep] <- r.fitted[TT-i.0]
			df$r1.Pr[i.rep] <- mod$Pr[TT-i.0]
			df$r1.est[i.rep] <- mean(b$r[,TT-i.0], na.rm=T)
			df$r1.est.se[i.rep] <- b$r.se[i.0]
			df$r1.u95[i.rep] <- b$r.upper95[TT-i.0]
			df$r1.u66[i.rep] <- b$r.upper66[TT-i.0]
			df$r1.l66[i.rep] <- b$r.lower66[TT-i.0]
			df$r1.l95[i.rep] <- b$r.lower95[TT-i.0]
			
			r.est <- colMeans(b$r, na.rm=T)
			t.max <- which(r.est == max(r.est, na.rm=T))[1]
			df$rmax[i.rep] <- r.fitted[t.max]
			df$rmax.est[i.rep] <- mean(b$r[,t.max], na.rm=T)
			df$rmax.u95[i.rep] <- b$r.upper95[t.max]
			df$rmax.u66[i.rep] <- b$r.upper66[t.max]
			df$rmax.l66[i.rep] <- b$r.lower66[t.max]
			df$rmax.l95[i.rep] <- b$r.lower95[t.max]
		
			t.min <- which(r.est == min(r.est, na.rm=T))[1]
			df$rmin[i.rep] <- r.fitted[t.min]
			df$rmin.est[i.rep] <- mean(b$r[,t.min], na.rm=T)
			df$rmin.u95[i.rep] <- b$r.upper95[t.min]
			df$rmin.u66[i.rep] <- b$r.upper66[t.min]
			df$rmin.l66[i.rep] <- b$r.lower66[t.min]
			df$rmin.l95[i.rep] <- b$r.lower95[t.min]
	
			write.csv(file=paste0("USA_counties_TRV.rev=",i.rev,"_",i.data,"_sr.fixed.min=",sr.fixed.min,"_27May20.csv"), df)
			
			ff <- fits[1:length(x),]
	
			ff$state_county <- i.county
			ff$date <- dat$date
			ff$type <- "x"
			ff$val <- x	
			f <- ff
			
			ff$state_county <- i.county
			ff$date <- dat$date
			ff$type <- "x.fitted"
			ff$val <- x.fitted
			f <- rbind(f,ff)
			
			ff$state_county <- i.county
			ff$date <- dat$date
			ff$type <- "r.fitted"
			ff$val <- r.fitted
			f <- rbind(f,ff)
	
			ff$state_county <- i.county
			ff$date <- dat$date
			ff$type <- "Pr.fitted"
			ff$val <- mod$Pr.fitted
			f <- rbind(f,ff)
	
			ff$state_county <- i.county
			ff$date <- dat$date
			ff$type <- "r.se"
			ff$val <- b$r.se
			f <- rbind(f,ff)
	
			ff$state_county <- i.county
			ff$date <- dat$date
			ff$type <- "r.u95"
			ff$val <- b$r.upper95
			f <- rbind(f,ff)
	
			ff$state_county <- i.county
			ff$date <- dat$date
			ff$type <- "r.u66"
			ff$val <- b$r.upper66
			f <- rbind(f,ff)
	
			ff$state_county <- i.county
			ff$date <- dat$date
			ff$type <- "r.l66"
			ff$val <- b$r.lower66
			f <- rbind(f,ff)
	
			ff$state_county <- i.county
			ff$date <- dat$date
			ff$type <- "r.l95"
			ff$val <- b$r.lower95
			f <- rbind(f,ff)
			
			if(start.flag){
				write.table(file=paste0("USA_counties_fits_TVR.rev=",i.rev,"_",i.data,"_sr.fixed.min=",sr.fixed.min,"_27May20.csv"), f, row.names=F, sep=",")
				start.flag <- F
			}else{
				write.table(file=paste0("USA_counties_fits_TVR.rev=",i.rev,"_",i.data,"_sr.fixed.min=",sr.fixed.min,"_27May20.csv"), f, row.names=F, sep=",", append = T, col.names = F)
			}
		}
	}	
	return("complete")
}

###########################################################
# compute r
###########################################################

d <- read.csv("USA_county_covid-19_data_26May20.csv", header=T)
d$date <- as.Date(d$date)
d$state_county <- as.factor(d$state_county)
d$state <- as.factor(d$state)
d$abbr <- as.factor(d$abbr)

fun <- function(X) {
	x <- list(list(data ="deaths", rev = F),
			list(data ="deaths", rev = T),
			list(data ="cases", rev = F),
			list(data ="cases", rev = T))[[X]]
	compute_r(x, d, annealing = T, se.fixed = 0, sr.fixed = NA, sr.fixed.min = 0.02, sr0.fixed = NA, sm.start = 0.5, sr.start = 0.04, delta = .5, k = 10, moving.ave = 1, nboot = 100)
}
registerDoParallel(cl = 4)
getDoParWorkers()
foreach(X=1:4, .verbose=T, .packages="mgcv") %dopar% fun(X)
registerDoSEQ()
	
```
