## Supplementary material for "Estimating and explaining the spread of COVID-19 at the county level in the USA": Source code and compiled data: tvr_27May20.R

```
#' Time-Varying Intrinsic Rate of Increase model
#'
#' Anthony R. Ives

####################################################
# Begin tvr

tvr <- function(X, rev = F, U = NULL, count.data = T, r0.start = NA, se.start = 0, sm.start = 0.01, sr.start = 0.01, se0.start = 0.01, sr0.start = 0.01, X0.start = NA, c.start = if (is.null(U)) NULL else array(0, dim = dim(U)[2]), r0.fixed = NA, se.fixed = 0, sm.fixed = NA, se0.fixed = 0, sr0.fixed = 0, X0.fixed = NA, sr.fixed = NA, c.fixed = if (is.null(U)) NULL else array(NA, dim = dim(U)[2]), annealing = T, Tsamplefract = .9, show.fig = F, method = "Nelder-Mead", maxit = 10^4, optim.control = NULL, maxit.SANN = 5, optim.SANN.control = NULL) {
  
  require("GenSA")
  
	X <- as.matrix(X)
  	Tmax <- nrow(X)

	if (!is.null(U)) {
		U <- as.matrix(U)
		nu <- ncol(U)
	}

  	if(rev == T) {
  		X <- as.matrix(X[Tmax:1])
  		if (!is.null(U)) U <- as.matrix(U[Tmax:1,])
  	}

	Tsample <- floor(Tsamplefract * Tmax)

	xx <- X[1:(Tsample - 1)]
	xx <- cbind(xx, rep(1, Tsample - 1))
	if (!is.null(U))
		xx <- cbind(xx, U[1:(Tsample - 1)])
	yy <- X[2:Tsample]

	# remove NAs for lm.fit
	zz <- cbind(yy, xx)
	xx <- xx[!rowSums(is.na(zz)),]
	yy <- yy[!rowSums(is.na(zz))]
	ar.init <- lm.fit(x = xx, y = yy)
	
	r0.init <- ar.init$coef[2]
	
	sm.init <- sm.start
	sr.init <- sr.start
	se.init <- se.start
	se0.init <- se0.start
	sr0.init <- sr0.start
	X0.init <- X[1]
	
	c.init <- c.start

	par.init <- c(r0.init, se.init, sm.init, sr.init, se0.init, sr0.init, X0.init, c.init)
	par.start <- c(r0.start, se.start, sm.start, sr.start, se0.start, sr0.start, X0.start, c.start)
	par.fixed <- c(r0.fixed, se.fixed, sm.fixed, sr.fixed, se0.fixed, sr0.fixed, X0.fixed, c.fixed)

	# set up variables for fitting
	par.full <- par.init
	par.full[!is.na(par.start)] <- par.start[!is.na(par.start)]

	par.full[!is.na(par.fixed)] <- par.fixed[!is.na(par.fixed)]
	par <- par.full[is.na(par.fixed)]

	if (is.null(optim.control)) 
		optim.control = list(maxit = maxit)
	if (is.null(optim.SANN.control)) 
		optim.SANN.control = list(temp = 1, tmax = 10, maxit = maxit.SANN)

	counter <- 1
	fitted.values <- NULL
	if (annealing == T) {
		if (!is.null(U)) {
			par.upper <- c(100, 10, 10, 10, 10, 10, max(X, na.rm=T)+1, array(100, dim = nu))
			par.lower <- c(-100, -10^-4, -10^-4, -10^-4, -10^-4, -10^-4, min(X, na.rm=T)-1, array(-100, dim = nu))
		} else {
			par.upper <- c(100, 10, 10, 10, 10, 10, max(X, na.rm=T)+1)
			par.lower <- c(-100, -10^-4, -10^-4, -10^-4, -10^-4, -10^-4, min(X, na.rm=T)-1)
		}

		par.upper <- par.upper[is.na(par.fixed)]
		par.lower <- par.lower[is.na(par.fixed)]

		optSANN <- GenSA(fn = tvr_ml, par = par, lower = par.lower, upper = par.upper, X = X, U = U, par.fixed = par.fixed, 
			count.data = count.data, control = list(smooth = F, maxit = maxit.SANN))
		par <- optSANN$par
	}

	opt <- optim(fn = tvr_ml, par = par, X = X, U = U, par.fixed = par.fixed, count.data = count.data, method = method, control = optim.control)

	# retrieve final fitted values
	fit <- tvr_fit(opt$par, X = X, U = U, par.fixed, count.data = count.data, show.fig = show.fig)

  	if(rev == T) {
  		X <- as.matrix(X[Tmax:1])
  		U <- if (!is.null(U)) as.matrix(U[Tmax:1,])
  		X.fitted = as.matrix(fit$X[Tmax:1])
  		r.fitted = -as.matrix(fit$r[Tmax:1])
  		Px.fitted = as.matrix(fit$Px[Tmax:1])
  		Pr.fitted = as.matrix(fit$Pr[Tmax:1])
  		me.fitted = as.matrix(fit$me[Tmax:1])
  	}else{
  		X.fitted = fit$X
  		r.fitted = fit$r
  		Px.fitted = fit$Px
  		Pr.fitted = fit$Pr
  		me.fitted = fit$me
  		
  	}
  	
	results <- list(X = X, rev = rev, U = U, count.data = count.data, 
					se = fit$se, sm = fit$sm, sr = fit$sr, se0 = fit$se0, sr0 = fit$sr0, 
					cc = fit$cc, X0 = fit$X0, r0 = fit$r0, 
					logLik = fit$logLik, AIC = fit$AIC, npar = fit$npar, 
					X.fitted = X.fitted, r.fitted = r.fitted, Px.fitted = Px.fitted, 
					Pr.fitted = Pr.fitted, me.fitted = fit$me,
					r0.start = r0.start, sr.start = sr.start, se.start = se.start, sm.start = sm.start, c.start = c.start, 
					r0.fixed = r0.fixed, se.fixed = se.fixed, sm.fixed = sm.fixed, X0.fixed = X0.fixed, 
					sr.fixed = sr.fixed, sr0.fixed = sr0.fixed, se0.fixed = se0.fixed, c.fixed = c.fixed, 
					opt.par = opt$par, par.full = par.full, par.fixed = par.fixed, 
					Tsamplefract = Tsamplefract, annealing = annealing, 
					optim.control = optim.control, optim.SANN.control = optim.SANN.control, convergence = opt$convergence)
  class(results) <- "tvr"
    
  return(results)
}
# End tvr
####################################################

####################################################
# Begin tvr_ml
tvr_ml <- function(par, X, U, par.fixed, count.data) {
	Tmax <- dim(X)[1]

	par.full <- par.fixed
	par.full[is.na(par.fixed)] <- par

	r0 <- par.full[1]
	s2e <- par.full[2]^2
	s2m <- par.full[3]^2
	s2r <- par.full[4]^2
	s2e0 <- par.full[5]^2
	s2r0 <- par.full[6]^2
	X0 <- par.full[7]
	if (!is.null(U)) {
		nu <- dim(U)[2]
		cc <- matrix(par.full[8:(7 + nu)], ncol = 1)
	}else{
		cc <- NULL
	}

	x <- X0
	r <- r0
	
	if(is.na(par.fixed[7])){
		S <- diag(c(s2e, s2r))
		if(is.na(par.fixed[5])) s2e0 <- s2e
		if(is.na(par.fixed[6])) s2r0 <- s2r
		PP <- diag(c(s2e0, s2r0))
	}else{
		S <- diag(c(0, s2r))
		PP <- diag(c(0, s2r0))
	}
	
	Z <- matrix(c(1,0), nrow=1)
	B <- matrix(c(1,0,1,1), 2,2)

	logFt <- 0
	vFv <- 0

	for (t in 1:Tmax) {

		# PREDICTION EQUATIONS
		if (t > 1) {
			PP <- B %*% PP %*% t(B) + S

			x <- r + x
			if (!is.null(U)) {
				if (ncol(U) == 1) {
					r <- r + as.numeric(U[t-1] * cc)
				} else {
					r <- r + as.numeric(U[t-1, ] %*% cc)
				}
			}
		}

		# UPDATING EQUATIONS
		if (!is.na(X[t])) {
			if (count.data == T) {
#				FF <- PP[1,1] + log(s2m * exp(-x) + 1)
				FF <- PP[1,1] + s2m + exp(-x)
			} else {
				FF <- PP[1,1] + s2m
			}
			invF <- 1/FF

			y <- matrix(c(x, r), ncol=1)
			v <- X[t] - Z %*% y

			y <- y + PP %*% t(Z) %*% invF %*% v
			PP <- PP - PP %*% t(Z) %*% invF %*% Z %*% PP

			x <- y[1]
			r <- y[2]

			# TERMS OF LIKELIHOOD FUNCTION
			if (!is.nan(FF) && FF > 0) {
				logFt <- logFt + log(FF)
			} else {
				logFt <- 10^10
			}

			vFv <- vFv + t(v) %*% invF %*% v
		}
	}

	LL <- logFt + vFv
	if (is.complex(LL)) 
		LL <- 10^10
	return(LL)
}
# End tvr_ml
####################################################


####################################################
# Begin tvr_fit
 tvr_fit <- function(par, X, U, par.fixed, count.data, show.fig) {

	Tmax <- dim(X)[1]

	par.full <- par.fixed
	par.full[is.na(par.fixed)] <- par

	r0 <- par.full[1]
	s2e <- par.full[2]^2
	s2m <- par.full[3]^2
	s2r <- par.full[4]^2
	s2e0 <- par.full[5]^2
	s2r0 <- par.full[6]^2
	X0 <- par.full[7]
	if (!is.null(U)) {
		nu <- dim(U)[2]
		cc <- matrix(par.full[8:(7 + nu)], ncol = 1)
	}else{
		cc <- NULL
	}

	x <- X0
	r <- r0
	
	if(is.na(par.fixed[7])){
		S <- diag(c(s2e, s2r))
		if(is.na(par.fixed[5])) s2e0 <- s2e
		if(is.na(par.fixed[6])) s2r0 <- s2r
		PP <- diag(c(s2e0, s2r0))
	}else{
		S <- diag(c(0, s2r))
		PP <- diag(c(0, s2r0))
	}
	
	Z <- matrix(c(1,0), nrow=1)
	B <- matrix(c(1,0,1,1), 2,2)

	X.fit <- array(NA, length(X))
	r.fit <- array(NA, length(X))
	Px.fit <- array(NA, length(X))
	Pr.fit <- array(NA, length(X))

	me.fit <- array(NA, length(X))

	logFt <- 0
	vFv <- 0

	for (t in 1:Tmax) {

		# PREDICTION EQUATIONS
		if (t > 1) {
			PP <- B %*% PP %*% t(B) + S

			x.old <- x
			x <- r + x
			if (!is.null(U)) {
				if (ncol(U) == 1) {
					r <- r + as.numeric(U[t-1] * cc)
				} else {
					r <- r + as.numeric(U[t-1, ] %*% cc)
				}
			}
		}

		# UPDATING EQUATIONS
		if (!is.na(X[t])) {
			if (count.data == T) {
#				FF <- PP[1,1] + log(s2m * exp(-x) + 1)
				FF <- PP[1,1] + s2m + exp(-x)
			} else {
				FF <- PP[1,1] + s2m
			}
			invF <- 1/FF

			y <- matrix(c(x, r), ncol=1)
			v <- X[t] - Z %*% y

			y <- y + PP %*% t(Z) %*% invF %*% v
			PP <- PP - PP %*% t(Z) %*% invF %*% Z %*% PP

			x <- y[1]
			r <- y[2]

			X.fit[t] <- x
			r.fit[t] <- r
			Px.fit[t] <- PP[1,1]
			Pr.fit[t] <- PP[2,2]
			me.fit[t] <- X[t] - x


			# TERMS OF LIKELIHOOD FUNCTION
			if (!is.nan(FF) && FF > 0) {
				logFt <- logFt + log(FF)
			} else {
				logFt <- 10^10
			}

			vFv <- vFv + t(v) %*% invF %*% v
		}
	}

	LL <- logFt + vFv
	if (is.complex(LL)) 
		LL <- 10^10

	npar <- length(par)
	logLik <- -((Tmax - 1)/2) * log(2 * pi) - LL/2

	npar <- length(par)
	logLik <- -((Tmax - 1)/2) * log(2 * pi) - LL/2
	AIC <- -2 * logLik + 2 * npar
	
	if (LL >= 10^10) {
		X.fit <- NULL
		r.fit <- NULL
	}

	if(show.fig){
		plot(1:Tmax, X/max(X, na.rm = T), typ = "p", xlab = "Time", ylab = "Y (o), Y.fitted (b), and r.fitted (r)", main = paste("logLik=", 
			0.001 * round(1000 * logLik)))
		lines(1:Tmax, X.fit/max(X, na.rm = T), col = "blue")
		lines(1:Tmax, r.fit/max(r.fit, na.rm = T), col = "red", pch = 20, cex = 0.5)
	
		if (!is.null(U)) {
			lines(1:Tmax, U * as.numeric(cc), col = "green")
		}		
	}
	
	return(list(r0 = r0, se = s2e^.5, sm = s2m^.5, sr = s2r^.5, se0 = s2e0^.5, sr0 = s2r0^.5, X0 = X0, cc = cc, logLik = logLik, AIC = AIC, npar = npar, X = X.fit, r = r.fit, Px = Px.fit, Pr = Pr.fit, me = me.fit))
}
# End tvr_fit
####################################################


####################################################
# Begin summary.tvr

summary.tvr <- function(x, ...) {
  
  cat("\nCall: tvr \n")
  
  cat("\nlogLik =", x$logLik)
  cat(",  AIC = ", x$AIC, " [df = ", x$npar, "]\n", sep = "")
  
  if(x$rev == F){
	cat("\nInitial value of r\n")
	cat("\tr0 = ", x$r0, "\n", sep = "")	
  } else{
 	cat("\nValue of r(Tmax) (at the start of the reversed time series)\n")
	cat("\tr0 = ", x$r0, "\n", sep = "")	
  } 	
  
  cat("\nVariation in r (as a standard deviation)\n")
  cat("\tsr = ", x$sr, "\n", sep = "")
  
  if (!is.null(x$cc)) {
  	if(x$rev == F){
    	cat("\n\nCoefficients for U\n")
    	for (i in 1:length(x$cc)) cat("\tc[", i - 1, "] = ", x$cc[i], "\n", sep = "")
    }else{
    	cat("\n\nCoefficients for U (in order of the original time series)\n")
    	for (i in 1:length(x$cc)) cat("\tc[", i - 1, "] = ", x$cc[length(x$cc) - i + 1], "\n", sep = "")
    }
  }
  cat("\nVariance parameters\n")
  cat("\tse = ", x$se, "\n", sep = "")
  cat("\tsm = ", x$sm, "\n", sep = "")
  
  cat("\nVariance in initial values\n")
  cat("\tse0 = ", x$se0, "\n", sep = "")
  cat("\tsr0 = ", x$sr0, "\n", sep = "")
  
  if(x$rev == F){
		cat("\nInitial value of X\n")
		cat("\tX0 = ", x$X0, "\n", sep = "")
	  if(!is.na(x$X0.fixed)){
	  	cat("\nInitial values of X assumed to be fixed with zero variance\n")  	
	  }else{
	  	cat("\nInitial values of X assumed to have standard error equal to se\n")
	  }
  }else{
		cat("\nValue of X at the end of the time series\n")
		cat("\tX0 = ", x$X0, "\n", sep = "")
	  if(!is.na(x$X0.fixed)){
	  	cat("\nValues of X at the end of the time series assumed to be fixed with zero variance\n")  	
	  }else{
	  	cat("\nValues of X at the end of the time series assumed to have standard error equal to se\n")
	  }
  }
  
  cat("\n")
}

print.tvr <- summary.tvr

# End summary.tvr
####################################################

####################################################
# Begin plot.tvr
plot.tvr <- function(x, par.envir = F, ...) {
  
  Tmax <- dim(x$X)[1]
  if(!is.null(x$U)) nrow.fig <- 3 else nrow.fig <- 2
  
  if(!par.envir) {
  	par.old <- par()
   	par(mfrow = c(nrow.fig, 1), mai = c(0.5, 1, .5, 0.1))
  }
  
  plot(1:Tmax, x$X, typ = "p", xlab = "", ylab = "X (o) and X.fitted (b)", main = paste("logLik=", round(x$logLik, digits = 3)))
  lines(1:Tmax, x$X.fitted, col = "blue")
  points(1:Tmax, x$X.fitted, col = "blue", pch = 20, cex = 0.5)
  plot(1:Tmax, x$r.fitted, xlab = "", ylab = "r.fitted (r)", col = "red", pch = 20, cex = 0.7, typ = "l")
  points(1:Tmax, x$r.fitted, col = "red", pch = 20, cex = 0.7)

  if (!is.null(x$U)) {
    U <- as.matrix(x$U)
    if (ncol(U) == 1) {
      plot(1:Tmax, U * as.numeric(x$cc), xlab = "", col = "green")
    } else {
      plot(1:Tmax, U[, 1] * as.numeric(x$cc[1]), xlab = "", col = "green")
      for (i in 2:ncol(U)) lines(1:Tmax, U[, i] * as.numeric(x$cc[i]), col = "green")
    }
  }
  if(!par.envir) par(mfrow = par.old$mfrow, mai = par.old$mai)

}
# End plot.tvr
####################################################


####################################################
# Begin simulate_tvr
simulate_tvr <- function(z, rev = F, r.order = c(1,0,0), count.data = T) {
	require("forecast")
	
	Tmax <- length(z$X.fitted)
	se <- z$se
	s2m <- z$sm^2
	cc <- z$cc
	
	U <- z$U
	cc <- z$cc

	r <- z$r.fitted
	X0 <- z$X0
	# reconstruct r
	if(rev == T){
		r <- -as.matrix(r[Tmax:1])
		X0 <- as.matrix(X0[Tmax:1])
	}	
	
	if(any(is.na(r))) r[is.na(r)] <- approx(r, xout = which(is.na(r)))$y

	r.dif <- r[-1] - r[-Tmax]
	if(!is.null(U)) r.dif <- r.dif - as.numeric(U[-Tmax,] %*% cc)
	
	r.dif <- r.dif - mean(r.dif)
	fit.dif <- Arima(r.dif, order=r.order)
	r.dif.sim <- simulate(fit.dif)
	if(!is.null(U)) r.dif.sim <- r.dif.sim + as.numeric(U[-Tmax,] %*% cc)
	
	r[1] <- r[1] + rnorm(n=1, sd = fit.dif$sigma2^.5)
	r[-1] <- r[-1] + (r.dif.sim - mean(r.dif.sim))/sqrt(2)

	x <- X0
	
	X <- array(NA, Tmax)
	X[1] <- X0

	for (t in 2:Tmax) {

		# PREDICTION EQUATIONS
		x <- r[t] + x + rnorm(n = 1, mean = 0, sd = se)
		
#		if(x <= log(.5)) x <- log(.5)
		X[t] <- x
	}
	
	# reconstruct em
	if (z$count.data == T) {
#		X <- X + rnorm(n = Tmax, sd = log(s2m * exp(-X) + 1)^.5)
#		X <- log(.5 + rpois(n=Tmax, lambda = exp(X + rnorm(n = Tmax, sd = s2m^.5))))
		X <- X + rnorm(n = Tmax, sd = (s2m + exp(-X))^.5)
	} else {
		X <- X + rnorm(n = Tmax, sd = s2m^.5)
	}

	if(rev == T){
		r <- -as.matrix(r[Tmax:1])
		X <- as.matrix(X[Tmax:1])
	}	

	return(list(X = X, r = r))
}
# End simulate_tvr
####################################################

####################################################
# Begin boot_trv
boot_tvr <- function(z, nboot = 10, r.resample = "ARIMA", r.order = c(1,0,0), me.resample = "ARIMA", me.order = c(1,0,0), annealing = z$annealing, replace = F, moving.ave = 1) {
	require("forecast")

	Tmax <- length(z$X.fitted)
	
	X.sim <- matrix(NA, nboot, Tmax)
	r.sim <- matrix(NA, nboot, Tmax)
	for(i.boot in 1:nboot) {
		sim <- simulate_tvr(z, 
			r.order = r.order)
		X.sim[i.boot,] <- sim$X
		r.sim[i.boot,] <- sim$r
	}

	X <- matrix(NA, nboot, Tmax)
	r <- matrix(NA, nboot, Tmax)
	nonconverg.count <- 0
	for(i.boot in 1:nboot) {
#		if(i.boot %% 10 == 0) show(i.boot)
		z.fit <- tvr(X.sim[i.boot,], rev = z$rev, U = z$U, sr.fixed = z$sr.fixed, 
				se.start = z$se, sm.start = z$sm, sr.start = z$sr,  
				annealing = annealing, show.fig = F)
		X[i.boot,] <- z.fit$X.fitted
		if(moving.ave == 1){
			r[i.boot,] <- z.fit$r.fitted
		}else{
			rr <- z.fit$r.fitted
			if(any(is.na(rr))) rr[is.na(rr)] <- approx(rr, xout = which(is.na(rr)))$y
			r[i.boot,] <- filter(rr, rep(1/moving.ave,moving.ave))
		}
		
		nonconverg.count <- nonconverg.count + z.fit$convergence
	}
	
	r0.est <- mean(r[,1], na.rm=T)
	r0.se <- sd(r[,1], na.rm=T)
	r0.bias <- r0.est - z$r.fitted[1]
	
	r1.est <- mean(r[,Tmax], na.rm=T)
	r1.se <- sd(r[,Tmax], na.rm=T)
	r1.bias <- r1.est - z$r.fitted[Tmax-1]
	
	r.upper66 <- array(NA, Tmax)
	r.lower66 <- array(NA, Tmax)
	for(t in 1:Tmax){
		r.upper66[t] <- sort(r[,t])[ceiling((1-.3333/2)*nboot)]
		r.lower66[t] <- sort(r[,t])[floor(.3333/2*nboot)]
	}

	if(nboot >= 0){
		r.upper95 <- array(NA, Tmax)
		r.lower95 <- array(NA, Tmax)
		for(t in 1:Tmax){
			r.upper95[t] <- sort(r[,t])[ceiling(.975*nboot)]
			r.lower95[t] <- sort(r[,t])[floor(.025*nboot)]
		}
	}else{
		r.upper95 <- NULL
		r.lower95 <- NULL
	}		
	
	r.se <- array(NA, Tmax)
	for(t in 1:Tmax){
		r.se[t] <- sd(r[,t])
	}
	
	
	return(list(X = X, U = z$U, r = r, X.sim = X.sim, r0.est = r0.est, r0.se = r0.se, r0.bias = r0.bias, r1.est = r1.est, r1.se = r1.se, r1.bias = r1.bias, r.upper66 = r.upper66, r.lower66 = r.lower66, r.upper95 = r.upper95, r.lower95 = r.lower95, r.se = r.se, nonconverg.count = nonconverg.count))
}
# End boot_trv
####################################################


####################################################
TVR_lag_fit <- function(mod, min.length, step){
	x <- mod$X
	mod.logLik <- mod
	
	range1 <- seq(from=min.length + 1, to=length(x) - min.length, by=step)
	logLik <- c(1,1,-10^10)

	for(lag1 in range1){
		if(lag1 >= (length(x) - 2*min.length)){
			U <- matrix(0, length(x), 1)
			U[lag1] <- 1
			mod0 <- tvr(x, U=U, se.fixed = mod$se.fixed, sr.fixed = mod$sr.fixed, 
				sr0.fixed = mod$sr0.fixed, se0.fixed = mod$se0.fixed,
				sm.start = mod$sm.start,  
				annealing = mod$annealing)

			if(logLik[3] < mod0$logLik) {
				show(c(lag1,mod0$logLik))
				plot(mod0)
				logLik <- c(lag1, NA, mod0$logLik)
				mod.logLik <- mod0
				U <- U
			}
		}else{
			range2 <- seq(from=lag1 + min.length , to=length(x) - min.length, by=step)
			for(lag2 in range2){
				U <- matrix(0, length(x), 2)
				U[lag1,1] <- 1
				U[lag2,2] <- 1
				mod0 <- tvr(x, U=U, se.fixed = mod$se.fixed, sr.fixed = mod$sr.fixed, 
					sr0.fixed = mod$sr0.fixed, se0.fixed = mod$se0.fixed,
					sm.start = mod$sm.start,  
					annealing = mod$annealing)
	
				if(logLik[3] < mod0$logLik) {
					show(c(lag1,lag2, mod0$logLik))
					plot(mod0)
					logLik <- c(lag1, lag2, mod0$logLik)
					mod.logLik <- mod0
					U <- U
				}
			}
		}
	}
	return(list(mod = mod.logLik, logLik=logLik, U=U))
}

# End TVR_lag_fit
###########################################################


###########################################################
# SIR simulated data

covid19_sim <- function(initx, I.init, b, surv = .98, se = .2, sm.incub = .2, sm.mort = .2, h.report = 0.5, show.fig = T) {
	
	duration <- 25
	Tmax <- length(b)
	
	# Transmission (Li)
	m <- 7.5
	v <- 3.4^2
	
	v/m^2
	shape <- 2.35
	gamma(1 + 2/shape)/gamma(1 + 1/shape)^2 - 1
	scale <- m/gamma(1 + 1/shape)
	
	pdf.trans <- dweibull(1:duration, shape=shape, scale = scale)
	pdf.trans <- pdf.trans/sum(pdf.trans)
	
	# Mortality (Li)
	m <- 18.5
	v <- 3.4^2
	
	v/m^2
	shape <- 10
	gamma(1 + 2/shape)/gamma(1 + 1/shape)^2 - 1
	scale <- m/gamma(1 + 1/shape)
	
	pdf.mort <- dweibull(1:duration, shape=shape, scale = scale)
	pdf.mort <- pdf.mort/sum(pdf.mort)

	# Incubation: Ferretti: lognormal mean 5.5 days, sd 2.2 days
	m <- 5.5
	v <- 2.2^2
	
	s2 <- log(v/m^2 + 1)
	mu <- log(m) - s2/2
	
	pdf.incub <- dlnorm(1:duration, meanlog = mu, sdlog = s2^.5)
	pdf.incub <- pdf.incub/sum(pdf.incub)
	

	# correction for lognormal environmental variation
	# b <- b/exp(se^2/2)
	
	B <- matrix(0,duration,duration)
	diag(B[-1,]) <- 1 - (1 - surv) * pdf.mort[1:(duration-1)]
	
	B[1,] <- b0 * pdf.trans * exp(se^2/2)
	
	x.init <- matrix(c(initx, rep(0,(duration - 1))), ncol=1)
	
	x1 <- matrix(c(1, rep(0,(duration - 1))), ncol=1)
	R <- NULL
	for(t in 1:duration){
		x1 <- B %*% x1
		R <- c(R, x1[1])
		x1[1] <- 0
	}
	R0 <- sum(R)
	
	r0 <- log(max(abs(eigen(B)$values)))
	
	# mean field simulation
	x <- x.init
	
	Ilist <- array(NA, Tmax)
	Ilist[1] <- sum(x)
	
	Dlist <- array(NA, Tmax)
	Dlist[1] <- 0
	
	rlist <- array(NA, Tmax)
	rlist[1] <- r0
	
	for(t in 2:Tmax){
		
		B[1,] <- b[t] * pdf.trans
		# correcting for lognormal
		B[1,] <- B[1,] * exp(se^2/2)
		D <- (1 - surv) * pdf.mort %*% x
		
		x <- B %*% x
	
		Ilist[t] <- pdf.incub %*% x
		Dlist[t] <- D
		rlist[t] <- log(max(abs(eigen(B)$values)))
	}
	
	
	X <- matrix(NA, 2, Tmax)
	x <- 0
	while(sum(x) == 0) x <- rpois(n = duration, lambda = I.init * x.init/sum(x.init))
	X[,1] <- c(sum(x), 0)
	x0 <- x
	
	r <- rep(NA, Tmax)
	r[1] <- r0
	
	for(t in 2:Tmax){
		
		B1 <- b[t] * pdf.trans * exp(rnorm(1, mean = 0, sd = se))
		D <- rbinom(n = duration, size = x, prob = (1 - surv) * pdf.mort * exp(rnorm(1,sd=sm.mort)))
		x <- x - D
		
		I.new <- rpois(n = 1, lambda = B1 %*% x)
		x[2:duration] <- x[1:(duration-1)]
		x[1] <- I.new
		
		C <- rbinom(n = duration, size = x, prob = h.report * pdf.incub * exp(rnorm(1,sd=sm.incub))) 
		X[,t] <- c(sum(C, na.rm=T), sum(D, na.rm=T))
		
		B[1,] <- B1
		r[t] <- log(max(abs(eigen(B)$values)))
	
	}
	
	if(show.fig){
		par(mfcol=c(2,2))
		plot(X[1,]/h.report, log="y", ylim=c(.1, max(X, na.rm = T)), col="green")
		points(X[2,])
		lines(Ilist * I.init/initx, col="green")
		lines(Dlist * I.init/initx)
		
		plot(rlist, typ="l", col="red")
		points(r, col="red")
		
		plot(X[1,]/h.report, col="green")
		lines(Ilist * I.init/initx, col="green")
		
		plot(X[2,])
		lines(Dlist * I.init/initx)
	}
	
	return(list(X = X, r = rlist, I = Ilist, D = Dlist, b = b))
}
```
